# Prehabilitative Exercise Mitigates Immobilization-Induced Atrophy Through Muscle Reserve Rather Than Resistance to Disuse: The PRE-EX Randomized Controlled Trial

**DOI:** 10.64898/2026.09.25.26363998

**Authors:** Casper Soendenbroe, Sara D. Pedersen, Leonard Pulevic, Amanda El Khoury, Helena L. Christiansen, Frederik H. Linden, Rene B. Svensson, S. Peter Magnusson, Peter Schjerling, Michael Kjaer, Abigail L. Mackey

**Author notes:** **Corresponding Authors:** Casper Soendenbroe, PhD, Abigail L. Mackey, PhD, DMSc.

## Abstract

Periods of limb immobilization are common in clinical care and lead to rapid losses of muscle mass and strength. We therefore examined whether prehabilitative exercise improves outcomes after disuse by increasing pre-immobilization muscle reserve or by attenuating the biological response to immobilization. Here, we report the primary and muscle-focused secondary outcomes of the PRE-EX (PREhabilitative EXercise) randomized controlled trial (ClinicalTrials.gov: NCT06205784). Fifty-four healthy younger and older women completed four weeks of supervised resistance exercise or no training before eight days of unilateral limb immobilization, followed by four weeks of rehabilitation. From baseline to pre-immobilization, prehabilitation increased quadriceps cross-sectional area (CSA) by 6.7%, maximal voluntary contraction (MVC) by 15.2%, and muscle fiber CSA by 17.7%. Immobilization subsequently reduced quadriceps CSA by 3.2% over 5 days and MVC by ∼20%, with similar relative losses in the Prehab and No-train groups. Consequently, quadriceps CSA, the study’s primary outcome, remained greater in Prehab than in No-train after immobilization and following rehabilitation, while MVC was greater following rehabilitation. Prehabilitation did not preserve markers of neuromuscular innervation or produce a detectable post-immobilization transcriptomic signature. Instead, immobilization elicited an extensive transcriptional response comprising more than 7000 differentially expressed genes, characterized by activation of inflammatory, proteolytic, stress, and denervation-associated programs together with suppression of mitochondrial and muscle-structural pathways. Network analysis further linked these coordinated molecular programs to disuse-related changes in muscle size, strength, and morphology. The effects of prehabilitation and the subsequent response to immobilization were similar in younger and older women. These findings demonstrate that short-term prehabilitative exercise mitigates the consequences of limb immobilization by establishing greater muscle and strength reserve before disuse, rather than by attenuating the physiological or molecular response to immobilization.

## Introduction

Periods of enforced limb disuse are common in clinical care, occurring after injury or surgery, during casting or bracing, or as part of prescribed activity restriction (*1*). Experimental models such as bed rest, casting and unilateral immobilization demonstrate rapid muscle loss of ∼0.5% per day, with strength declining two- to four-fold faster over the same interval (*2–4*). Although subsequent exercise-based rehabilitation can partially restore functional capacity, recovery is often incomplete and slower in older adults (*5–11*), underscoring the need for strategies to improve outcomes after predictable periods of disuse. In such settings, the physiological stress imposed by many operative procedures may also limit the capacity to respond to training post-operatively, positioning prehabilitation as a potentially important component of preventive care strategies (*12*). Importantly, women remain underrepresented in mechanistic experimental studies of disuse (*4*).

Prehabilitative exercise, defined as targeted physical conditioning delivered before a planned physiological stress or period of disuse, has shown promise across several clinical contexts (*13*), with reports of improved functional status in colorectal cancer, major abdominal surgery, and hip or knee arthroplasty (*14–20*). However, mechanistic insight remains limited, as existing trials are heterogeneous in design and outcomes, and do not resolve whether prehabilitation improves subsequent outcomes by increasing physiological reserve before disuse, attenuating the biological response during disuse, accelerating recovery afterwards, or through some combination of these mechanisms. Notably, the few mechanistic human studies examining pre-immobilization exercise have largely focused on brief interventions with short-term molecular readouts, leaving longer-term functional protection unresolved (*21, 22*).

A key unresolved issue is whether prehabilitation can influence the neuromuscular mechanisms underlying the disproportionate loss of strength relative to muscle atrophy. During short-term disuse, strength declines exceed losses of muscle mass by roughly two- to four-fold (*2, 3*), implicating mechanisms beyond muscle atrophy. Experimental work points to disturbances of muscle innervation - including increased expression of denervation-associated genes (*23*), greater prevalence of NCAM-positive fibers (*24*), circulating biomarkers of denervation–reinnervation (*25*), and motor unit remodeling (*26*) - as plausible contributors to disproportionate weakness. Importantly, both we and others have shown that acute and long-term exercise can influence markers of muscle innervation (*27–29*), raising the possibility that prehabilitation may act not only by increasing contractile tissue before disuse but also by preserving neuromuscular integrity during immobilization. Whether such preservation occurs in humans, and whether it relates to trajectories of muscle mass and strength during and after immobilization, remains unknown.

In this study, we tested if a short resistance-training-based prehabilitation intervention could alter trajectories of muscle mass, strength, and indices of neuromuscular innervation across a brief period of limb immobilization. We hypothesized that prehabilitation would; (1) increase physiological reserve before disuse, and (2) accelerate post-immobilization recovery of muscle mass and strength, potentially through preserved neuromuscular innervation, with age modifying the responses to both prehabilitation and immobilization. To test this, we conducted the PRE-EX (PREhabilitative EXercise) randomized controlled trial in healthy younger and older women, in which four weeks of supervised resistance exercise preceded eight days of unilateral limb immobilization followed by four weeks of rehabilitation. Here we report the primary outcome of quadriceps cross-sectional area (CSA) together with muscle-focused secondary outcomes including maximal voluntary contraction, muscle fiber size, denervation markers, and muscle RNA sequencing.

## Results

### Participants and trial conduct

From 417 individuals assessed for eligibility, 64 were included (31 younger, 33 older). Within each age group participants were randomized to No-Train or Prehab (Young No-Train n=14, Young Prehab n=17, Older No-Train n=17, Older Prehab n=16). Dropouts occurred in Young No-Train (n=1), Young Prehab (n=4), Older No-Train (n=3), and Older Prehab (n=2), yielding 13/13/14/14 participants, respectively. All analyses presented below were conducted in the per-protocol population (n=54), comprising participants who completed the study (Supplementary Material 1). Throughout, Group refers to Prehab versus No-Train, Age to younger versus older participants, Leg to the immobilized versus contralateral control leg, and Time to baseline, pre-immobilization/post-prehabilitation, post-immobilization, and post-rehabilitation.

Baseline characteristics of the per-protocol population are summarized in Table 1 (Data sheet 1). As expected, older women exhibited lower chair-stand performance and handgrip strength, together with higher HbA1c and CRP levels. Within each age group, Prehab and No-Train participants were well matched, with only a borderline lower inactive time in Prehab (p=0.044). No participants fell below European Working Group on Sarcopenia in Older People 2 reference thresholds for muscle strength, physical performance, or muscle quantity at baseline (*30*). Chronic disease and regular medication use were reported by approximately one-third to one-half of participants, without significant differences between age or intervention groups.

**Table 1.**
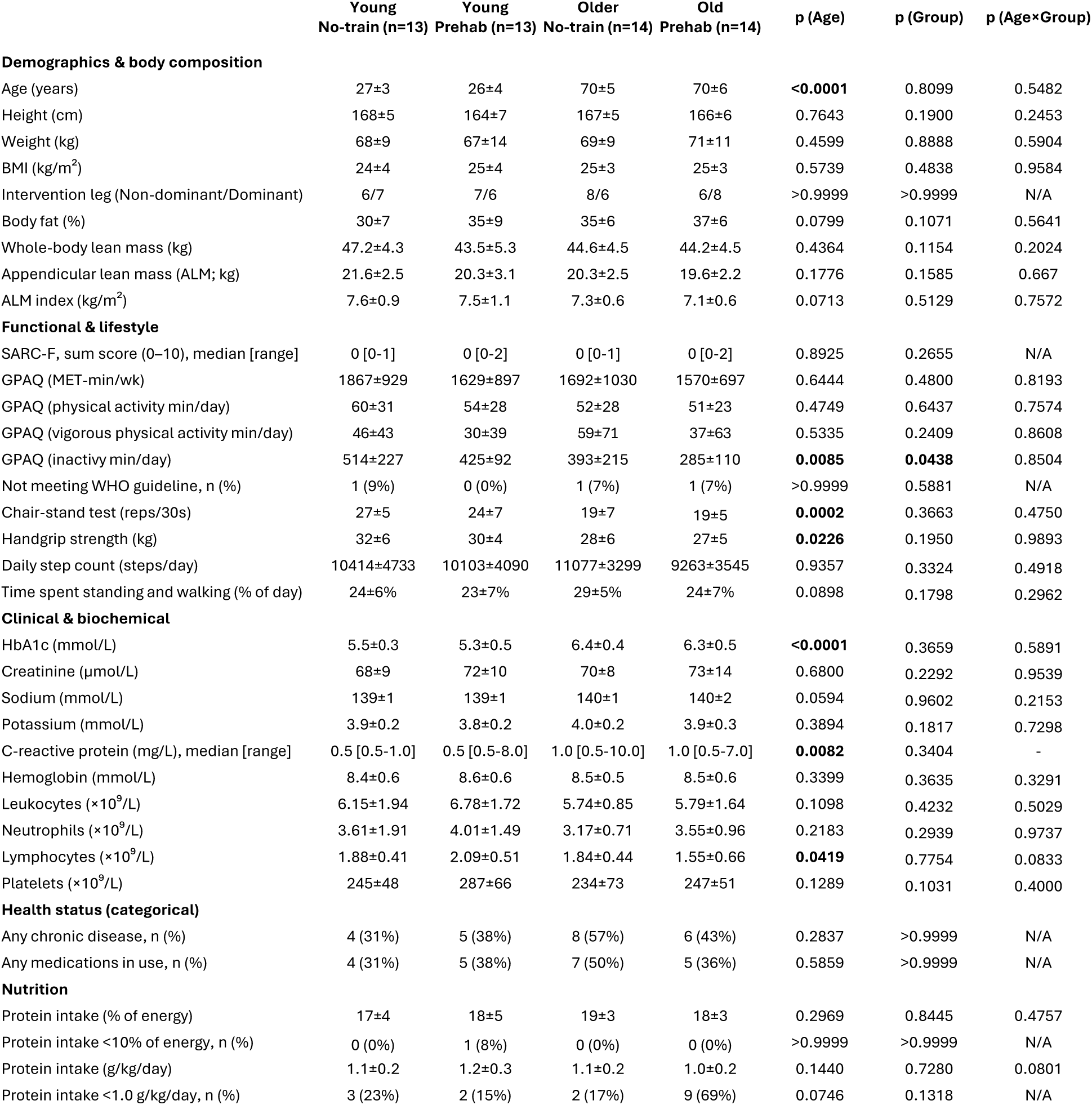
Baseline participant characteristics. Values are mean ± SD unless otherwise stated; skewed variables SARC-F, CRP) are reported as median [range]. For approximately normally distributed continuous variables, p values were derived from two-way ANOVA with factors Age (young vs older) and Group (Prehab vs Control) including the Age×Group interaction. Categorical variables were analysed using Fisher’s exact test on collapsed 2×2 tables to obtain p(Age) and p(Group). Clearly non-normal continuous variables (SARC-F, CRP) were analysed using Mann– Whitney tests on collapsed groups. The assay lower limit of quantification for CRP was 1 mg/L□¹. Values reported as “<1 mg/L□¹” were set to 0.5 mg/L for descriptive statistics and display only. Abbreviations: BMI, body mass index; GPAQ, Global Physical Activity Questionnaire; MET, metabolic equivalent; WHO, World Health Organization; HbA1c, glycated hemoglobin.

|  | Young<br>No-train (n=13) | Young<br>Prehab (n=13) | Older<br>No-train (n=14) | Old<br>Prehab (n=14) | p (Age) | p (Group) | p (Age×Group) |
| --- | --- | --- | --- | --- | --- | --- | --- |
| **Demographics & body composition** |  |  |  |  |  |  |  |
| Age (years) | 27±3 | 26±4 | 70±5 | 70±6 | <0.0001 | 0.8099 | 0.5482 |
| Height (cm) | 168±5 | 164±7 | 167±5 | 166±6 | 0.7643 | 0.1900 | 0.2453 |
| Weight (kg) | 68±9 | 67±14 | 69±9 | 71±11 | 0.4599 | 0.8888 | 0.5904 |
| BMI (kg/m<sup>2</sup>) | 24±4 | 25±4 | 25±3 | 25±3 | 0.5739 | 0.4838 | 0.9584 |
| Intervention leg (Non-dominant/Dominant) | 6/7 | 7/6 | 8/6 | 6/8 | >0.9999 | >0.9999 | N/A |
| Body fat (%) | 30±7 | 35±9 | 35±6 | 37±6 | 0.0799 | 0.1071 | 0.5641 |
| Whole-body lean mass (kg) | 47.2±4.3 | 43.5±5.3 | 44.6±4.5 | 44.2±4.5 | 0.4364 | 0.1154 | 0.2024 |
| Appendicular lean mass (ALM; kg) | 21.6±2.5 | 20.3±3.1 | 20.3±2.5 | 19.6±2.2 | 0.1776 | 0.1585 | 0.667 |
| ALM index (kg/m<sup>2</sup>) | 7.6±0.9 | 7.5±1.1 | 7.3±0.6 | 7.1±0.6 | 0.0713 | 0.5129 | 0.7572 |
| **Functional & lifestyle** |  |  |  |  |  |  |  |
| SARC-F, sum score (0–10), median [range] | 0 [0-1] | 0 [0-2] | 0 [0-1] | 0 [0-2] | 0.8925 | 0.2655 | N/A |
| GPAQ (MET-min/wk) | 1867±929 | 1629±897 | 1692±1030 | 1570±697 | 0.6444 | 0.4800 | 0.8193 |
| GPAQ (physical activity min/day) | 60±31 | 54±28 | 52±28 | 51±23 | 0.4749 | 0.6437 | 0.7574 |
| GPAQ (vigorous physical activity min/day) | 46±43 | 30±39 | 59±71 | 37±63 | 0.5335 | 0.2409 | 0.8608 |
| GPAQ (inactivity min/day) | 514±227 | 425±92 | 393±215 | 285±110 | **0.0085** | **0.0438** | 0.8504 |
| Not meeting WHO guideline, n (%) | 1 (9%) | 0 (0%) | 1 (7%) | 1 (7%) | >0.9999 | 0.5881 | N/A |
| Chair-stand test (reps/30s) | 27±5 | 24±7 | 19±7 | 19±5 | **0.0002** | 0.3663 | 0.4750 |
| Handgrip strength (kg) | 32±6 | 30±4 | 28±6 | 27±5 | **0.0226** | 0.1950 | 0.9893 |
| Daily step count (steps/day) | 10414±4733 | 10103±4090 | 11077±3299 | 9263±3545 | 0.9357 | 0.3324 | 0.4918 |
| Time spent standing and walking (% of day) | 24±6% | 23±7% | 29±5% | 24±7% | 0.0898 | 0.1798 | 0.2962 |
| **Clinical & biochemical** |  |  |  |  |  |  |  |
| HbA1c (mmol/L) | 5.5±0.3 | 5.3±0.5 | 6.4±0.4 | 6.3±0.5 | <0.0001 | 0.3659 | 0.5891 |
| Creatinine (μmol/L) | 68±9 | 72±10 | 70±8 | 73±14 | 0.6800 | 0.2292 | 0.9539 |
| Sodium (mmol/L) | 139±1 | 139±1 | 140±1 | 140±2 | 0.0594 | 0.9602 | 0.2153 |
| Potassium (mmol/L) | 3.9±0.2 | 3.8±0.2 | 4.0±0.2 | 3.9±0.3 | 0.3894 | 0.1817 | 0.7298 |
| C-reactive protein (mg/L), median [range] | 0.5 [0.5-1.0] | 0.5 [0.5-8.0] | 1.0 [0.5-10.0] | 1.0 [0.5-7.0] | **0.0082** | 0.3404 | - |
| Hemoglobin (mmol/L) | 8.4±0.6 | 8.6±0.6 | 8.5±0.5 | 8.5±0.6 | 0.3399 | 0.3635 | 0.3291 |
| Leukocytes (×10<sup>9</sup>/L) | 6.15±1.94 | 6.78±1.72 | 5.74±0.85 | 5.79±1.64 | 0.1098 | 0.4232 | 0.5029 |
| Neutrophils (×10<sup>9</sup>/L) | 3.61±1.91 | 4.01±1.49 | 3.17±0.71 | 3.55±0.96 | 0.2183 | 0.2939 | 0.9737 |
| Lymphocytes (×10<sup>9</sup>/L) | 1.88±0.41 | 2.09±0.51 | 1.84±0.44 | 1.55±0.66 | **0.0419** | 0.7754 | 0.0833 |
| Platelets (×10<sup>9</sup>/L) | 245±48 | 287±66 | 234±73 | 247±51 | 0.1289 | 0.1031 | 0.4000 |
| **Health status (categorical)** |  |  |  |  |  |  |  |
| Any chronic disease, n (%) | 4 (31%) | 5 (38%) | 8 (57%) | 6 (43%) | 0.2837 | >0.9999 | N/A |
| Any medications in use, n (%) | 4 (31%) | 5 (38%) | 7 (50%) | 5 (36%) | 0.5859 | >0.9999 | N/A |
| **Nutrition** |  |  |  |  |  |  |  |
| Protein intake (% of energy) | 17±4 | 18±5 | 19±3 | 18±3 | 0.2969 | 0.8445 | 0.4757 |
| Protein intake <10% of energy, n (%) | 0 (0%) | 1 (8%) | 0 (0%) | 0 (0%) | >0.9999 | >0.9999 | N/A |
| Protein intake (g/kg/day) | 1.1±0.2 | 1.2±0.3 | 1.1±0.2 | 1.0±0.2 | 0.1440 | 0.7280 | 0.0801 |
| Protein intake <1.0 g/kg/day, n (%) | 3 (23%) | 2 (15%) | 2 (17%) | 9 (69%) | 0.0746 | 0.1318 | N/A |

Baseline muscle characteristics differed between age groups, with older women showing lower quadriceps CSA, MVC, specific strength, and type II fCSA, together with greater type II SFI (Supplementary Material 2; Data sheet 1).

An overview of the study design is shown in Fig. 1A. Wearing the knee brace markedly reduced habitual activity (Supplementary Material 3A-B; Data sheet 1). Daily step count and time spent walking fell substantially during immobilization (Immobilization effect, p<0.0001 for all), confirming protocol adherence. Routine blood biomarkers showed no significant changes over time (Supplementary Material 4A-D). All participants completed at least 9 out of 10 scheduled exercise sessions in each training phase to which they were assigned.

**Figure 1.**
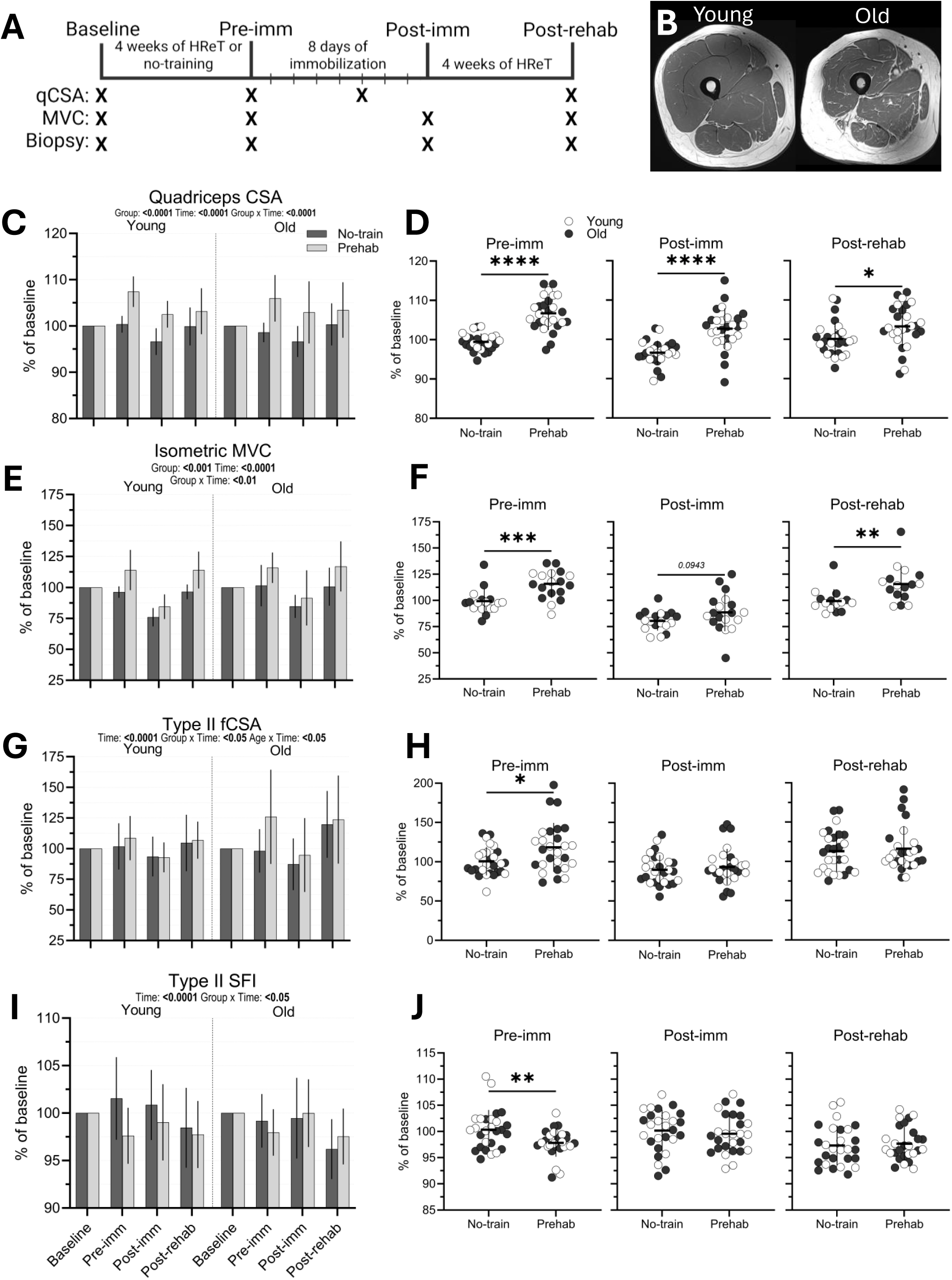
Prehabilitation builds muscle reserve before immobilization and improves muscle size and strength after disuse. (A) Study design. (B) Representative magnetic resonance imaging (MRI) images illustrating quadriceps cross-sectional area (CSA) in younger and older participants. (C-D) Quadriceps CSA of the immobilized leg, the primary outcome, measured by MRI (n=13-14 per group). (E-F) Maximal voluntary contraction (MVC) of the knee extensors measured isometrically using the same dynamometer at all time points (n=8-11 per group). Analyses including MVC measurements obtained using both dynamometers are presented in Supplementary Material 6. (G-H) Type II muscle fiber cross-sectional area (fCSA; n=13-14 per group). (I-J) Type II muscle fiber shape factor index (SFI; n=13-14 per group). Panels C, E, G, and I show values across the four study time points expressed as percentage of baseline. Panels D, F, H, and J show selected comparisons between the No-train and Prehab groups at the pre-immobilization, post-immobilization, and post-rehabilitation time points. Longitudinal outcomes were analyzed using linear mixed-effects models with Time, Age (younger vs older), and Group (Prehab vs No-train) as fixed factors, with model results reported in panels C, E, G, and I. Selected between-group comparisons were performed using unpaired t tests; significance for these comparisons is indicated in panels D, F, H, and J as *p<0.05, **p<0.01, ***p<0.001, and ****p<0.0001. Values are shown as mean ± SD, with individual values displayed in panels D, F, H, and J. Abbreviations: CSA, cross-sectional area; fCSA, muscle fiber cross-sectional area; MRI, magnetic resonance imaging; MVC, maximal voluntary contraction; SFI, shape factor index.

Adverse events were evaluated in all participants. Events were predominantly mild and transient, and none raised concerns regarding overall procedural safety. A small number of participants withdrew due to medical findings or procedure-related discomfort. Seasonal illnesses and minor musculoskeletal or skin reactions during immobilization and/or training were observed but self-limiting. One hospitalization occurred during immobilization (unrelated to intervention), and one fall potentially related to immobilization resulted in minor bruising only. No thrombotic events were observed. Detailed adverse event narratives are provided in Supplementary Material 5.

### Prehabilitative exercise increases skeletal muscle reserve and improves functional outcomes following immobilization

Quadriceps CSA, the primary outcome of the study, was assessed in the immobilized leg at baseline, pre-immobilization, 5 days into immobilization, and post-rehabilitation (Fig. 1B-D; Data sheet 1). A linear mixed-effects model revealed significant effects of Time and Group and a Time×Group interaction (all p<0.0001; Fig. 1C), with no effects involving Age. The magnitude of quadriceps CSA loss during immobilization, calculated as the percentage change from pre- to post-immobilization, was 3.2 ± 3.4% across participants, corresponding to approximately 0.6% per day. Selected comparisons between the No-train and

Prehab groups were subsequently performed at the pre-immobilization, post-immobilization, and post-rehabilitation time points (Fig. 1D). Following the 4-week prehabilitation intervention, the increase in quadriceps CSA from baseline was greater in Prehab than in No-train (6.8 %, p<0.0001), demonstrating that prehabilitative exercise increased muscle reserve before immobilization. The change from baseline remained greater in Prehab after immobilization (5.9 %, p<0.0001) and following rehabilitation (3.1 %, p<0.05), despite similar relative losses during immobilization.

Maximal voluntary contraction (MVC) of the knee extensors was assessed in the immobilized leg at baseline, pre-immobilization, post-immobilization, and post-rehabilitation using isometric testing (Fig. 1E–F). Owing to equipment malfunction, the primary MVC analysis was restricted to participants tested on the same dynamometer at all time points (n=37). Analyses including all available measurements are presented in Supplementary Material 6A–B (Data sheet 1). A linear mixed-effects model revealed significant effects of Time and Group, as well as a Time × Group interaction (all p<0.01), with no significant effects involving Age. Selected comparisons showed that the change in MVC from baseline was greater in Prehab than in No-train following the prehabilitation period (14.3%, p<0.001). This between-group difference in change from baseline was not statistically significant after immobilization (p=0.0943), but re-emerged following rehabilitation, when the change in MVC from baseline was again greater in Prehab than in No-train (14.0%, p<0.01).

Muscle fiber cross-sectional area (fCSA) was assessed by immunofluorescence staining. Type I fCSA showed only a main effect of Time (p<0.001), reflecting reductions in fiber size during immobilization across groups (Supplementary Material 7A; Data sheet 1). In contrast, type II fCSA exhibited significant effects of Time, Group×Time and Age×Time (all p<0.05; Fig. 1G-H). Selected comparisons showed that the change in type II fCSA from baseline was greater in Prehab than in No-train following the prehabilitation period (15.1%, p<0.05), whereas no significant between-group differences in change from baseline were observed after immobilization or rehabilitation. Myofiber shape was quantified using the shape factor index (SFI). Type I SFI remained unchanged throughout the study (Supplementary Material 7B), whereas type II SFI showed significant Time and Group×Time effects (p<0.05; Fig. 1I-J). Selected comparisons showed a greater reduction in type II SFI from baseline in Prehab than in No-train following the prehabilitation period (-2.6%, p<0.01), with no significant between-group differences in change from baseline thereafter. Fiber type distribution did not differ between groups or across time (Supplementary Material 7C).

Collectively, these findings indicate that prehabilitation increased muscle size and strength before immobilization, thereby improving outcomes after disuse, while the magnitude of immobilization-induced decline appeared similar between groups. Older women responded to prehabilitation and recovered following immobilization with no statistically significant evidence of a different response compared with younger women. We next examined whether the benefits of prehabilitation were accompanied by preservation of neuromuscular innervation.

### No protective effect of prehabilitative exercise on muscle innervation during disuse

Based on our previous observations that markers of neuromuscular innervation remodel in response to both acute (*27*) and long-term resistance training (*28*), and that prior exercise may imprint muscle cells in ways that support motor neuron survival (*31*), we investigated whether prehabilitation protected against immobilization-induced disruption of muscle innervation.

We first assessed the expression of two acetylcholine receptor (AChR) subunits, CHRNA1 and CHRND, by RT-qPCR, as both have previously been shown to increase substantially with disuse (*23*) (Fig. 2A–B; Data sheet 1). Both transcripts showed a robust main effect of Time (p < 0.0001), increasing approximately 6–8-fold following immobilization. This response was consistent with denervation-associated transcriptional remodeling during short-term disuse. No significant differences were observed between the No-train and Prehab groups.

**Figure 2.**
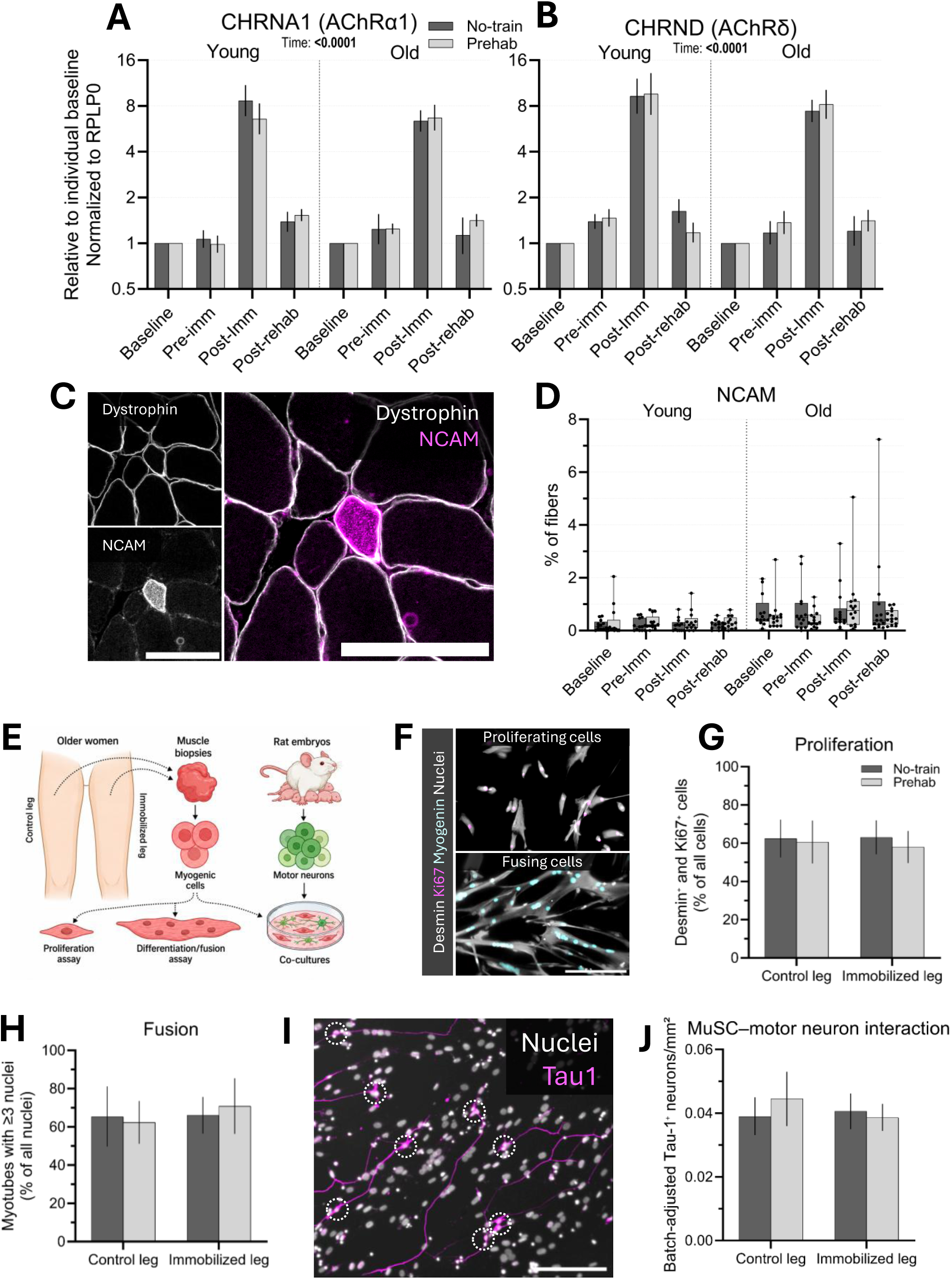
Immobilization induces denervation-associated transcriptional remodeling without evidence of neuromuscular protection by prehabilitation. (A–B) CHRNA1 and CHRND gene expression measured by RT-qPCR at baseline, pre-immobilization, post-immobilization, and post-rehabilitation (n=54). D Data are shown as geometric means with back-transformed SE calculated on the log2 scale, normalized to RPLP0 and expressed relative to individual baseline values. Statistical analyses on log-transformed data were performed using mixed-effects models with Time, Age, and Group as fixed factors. (C) Representative image of an NCAM-positive muscle fiber (scale bar = 100 µm). (D) NCAM-positive fibers expressed as a percentage of all analyzed fibers at baseline, pre-immobilization, post-immobilization, and post-rehabilitation (n=54). Data are shown as box plots with the median and interquartile range, whiskers indicating the minimum and maximum values, and individual data points overlaid. Changes over time were evaluated separately within each group using the Friedman test. (E) Schematic overview of the primary MuSC monoculture and MuSC–motor neuron co-culture experiments. (F) Representative images of proliferating and fusing MuSC cultures (scale bar = 100 µm). (G) Proliferation index, defined as the proportion of Desmin-positive cells co-expressing Ki67 (n=19). (H) Fusion index, defined as the proportion of nuclei contained within myotubes with ≥3 nuclei (n=18). (I) Representative image of a MuSC–motor neuron co-culture (scale bar = 200 µm); dashed circles indicate motor neurons. (J) Motor neuron abundance in MuSC–motor neuron co-culture (n=18). Values are shown as batch-adjusted Tau-1 neurons/mm² and were adjusted for between-experiment variation by subtracting the mean of the corresponding experimental batch and adding the global mean across all six experiments. Data in panels G, H, and J are shown as mean ± SD. Statistical analyses were performed using mixed-effects models with Leg and Group as fixed factors. Abbreviations: CHRNA1, cholinergic receptor nicotinic alpha 1 subunit; CHRND, cholinergic receptor nicotinic delta subunit; MuSC, muscle stem cell; NCAM, neural cell adhesion molecule; RT-qPCR, reverse-transcription quantitative polymerase chain reaction.

Muscle fiber denervation was next evaluated using NCAM immunofluorescence (Fig. 2C-D). The proportion of NCAM fibers showed substantial intra-individual variability and a non-normal distribution and was therefore analyzed using non-parametric statistics. No significant within-group changes were detected at any time point. At baseline, however, the proportion of NCAM fibers was greater in older than in younger participants (p<0.0001; Supplementary Material 7D), consistent with established age-related differences in neuromuscular integrity (*27, 32, 33*).

To determine whether immobilization or prehabilitation induced persistent changes in intrinsic muscle stem cell (MuSC) properties, proliferative and fusion capacities were assessed *in vitro* (Fig. 2E–F). Neither proliferative capacity (Fig. 2G) nor fusion capacity (Fig. 2H) differed between legs or intervention groups. MuSC–motor neuron interactions were then evaluated using a co-culture system with rat embryonic motor neurons (Fig. 2I–J). A trend toward a Leg×Group interaction was observed, suggesting a possible, but inconclusive, influence of prehabilitation on MuSC–motor neuron interactions.

Collectively, immobilization induced a marked increase in denervation-associated AChR transcripts, without detectable changes in NCAM-positive fibers or clear alterations in muscle stem cell–motor neuron interactions. Across these measures, there was no convincing evidence that prehabilitation preserved neuromuscular innervation. We therefore next examined whether its beneficial effects were accompanied by broader molecular protection against disuse, as would be expected if prehabilitation increased the intrinsic resistance of skeletal muscle to immobilization.

### Prehabilitation does not alter the muscle transcriptomic response to disuse

Given the absence of effects of prehabilitation on neuromuscular innervation, we used an unbiased transcriptomic approach to assess whether its physiological benefits were accompanied by broader molecular protection during disuse. Bulk RNA sequencing was performed on muscle biopsies obtained from both the control and immobilized legs at post-immobilization in all participants (Fig. 3A), thereby characterizing the post-immobilization molecular state associated with the immobilization phenotype. This design enabled simultaneous evaluation of the effects of Age (young vs older), Leg (control vs immobilized), and Group (No-Train vs Prehab). Following quality-control filtering, an average of 26910 genes were detected per sample.

**Figure 3.**
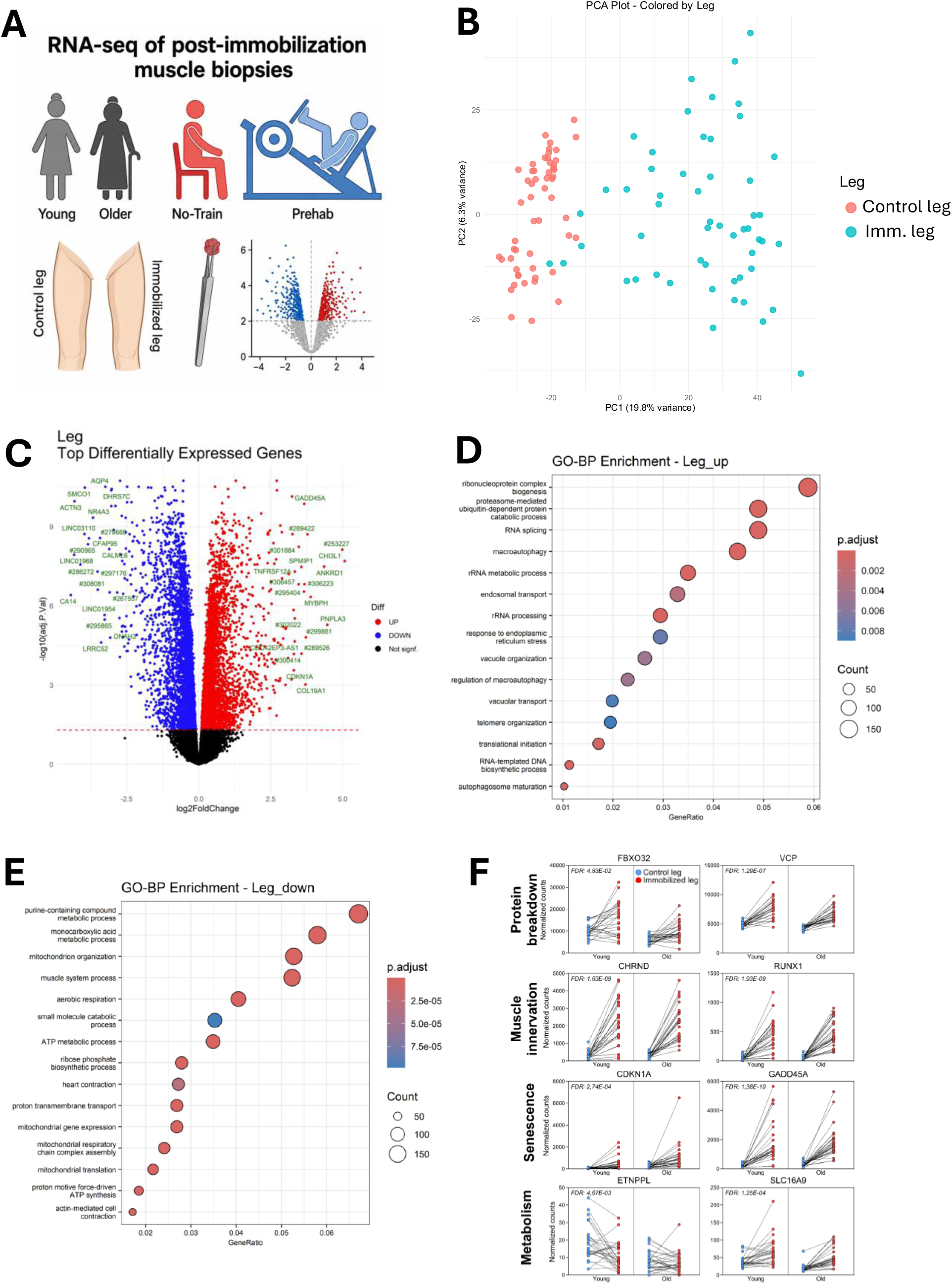
Limb immobilization induces extensive transcriptional remodeling that is not altered by prehabilitation. (A) Bulk RNA sequencing was performed on post-immobilization muscle biopsies obtained from both the control and immobilized legs of all participants (n=54). The schematic summarizes the transcriptomic analyses across Age (younger vs older), Group (No-train vs Prehab), and Leg (control vs immobilized). (B) Principal component analysis of normalized gene expression, showing separation by Leg along PC1 and by Age along PC2, with no distinct clustering by Group. (C) Differential expression analysis identified 7796 genes associated with Leg (FDR<0.05). (D–E) Gene Ontology biological-process enrichment analyses illustrating immobilization-induced transcriptional remodeling. (F) Expression of selected genes related to protein breakdown, denervation-associated remodeling, cellular stress, and metabolism. Differential expression analyses were corrected for multiple testing using the false discovery ratF3e, with significance defined as FDR<0.05. Analyses restricted to the No-train group are presented in Supplementary material 8D.

Principal component analysis revealed clear structure in the dataset (Supplementary Material 8A). Samples separated strongly by Leg along PC1 and by Age along PC2 and PC3 (Fig. 3B; Supplementary Material 8B; Data sheet 2), whereas Group did not form distinct clusters. Consistent with this, no differential gene expression attributable to prehabilitation was observed. Despite its clear effects on muscle size and strength, prehabilitation therefore did not produce a detectable post-immobilization transcriptomic signature. These findings support the interpretation that its benefits arose primarily from the greater muscle and strength reserve established before disuse, rather than from broad attenuation of the molecular response to immobilization.

### Disuse and ageing induce transcriptional remodeling in human skeletal muscle

Having found no broad transcriptomic effect of prehabilitation, we next characterized the molecular response to immobilization itself. This allowed us to determine whether immobilization-associated biological processes remained evident despite the benefits of prehabilitation. We also compared gene-expression signatures between younger and older participants to distinguish age-associated transcriptional differences from the response to disuse. Consistent with the patterns observed in the principal component analysis, differential expression analyses revealed pronounced effects of both Leg and Age. Immobilization was associated with 7796 differentially expressed genes (DEGs; 41.3 % of detected genes), whereas Age was associated with 1954 DEGs (10.3 % of detected genes; FDR < 0.05) (Fig. 3C and 4A).

**Figure 4.**
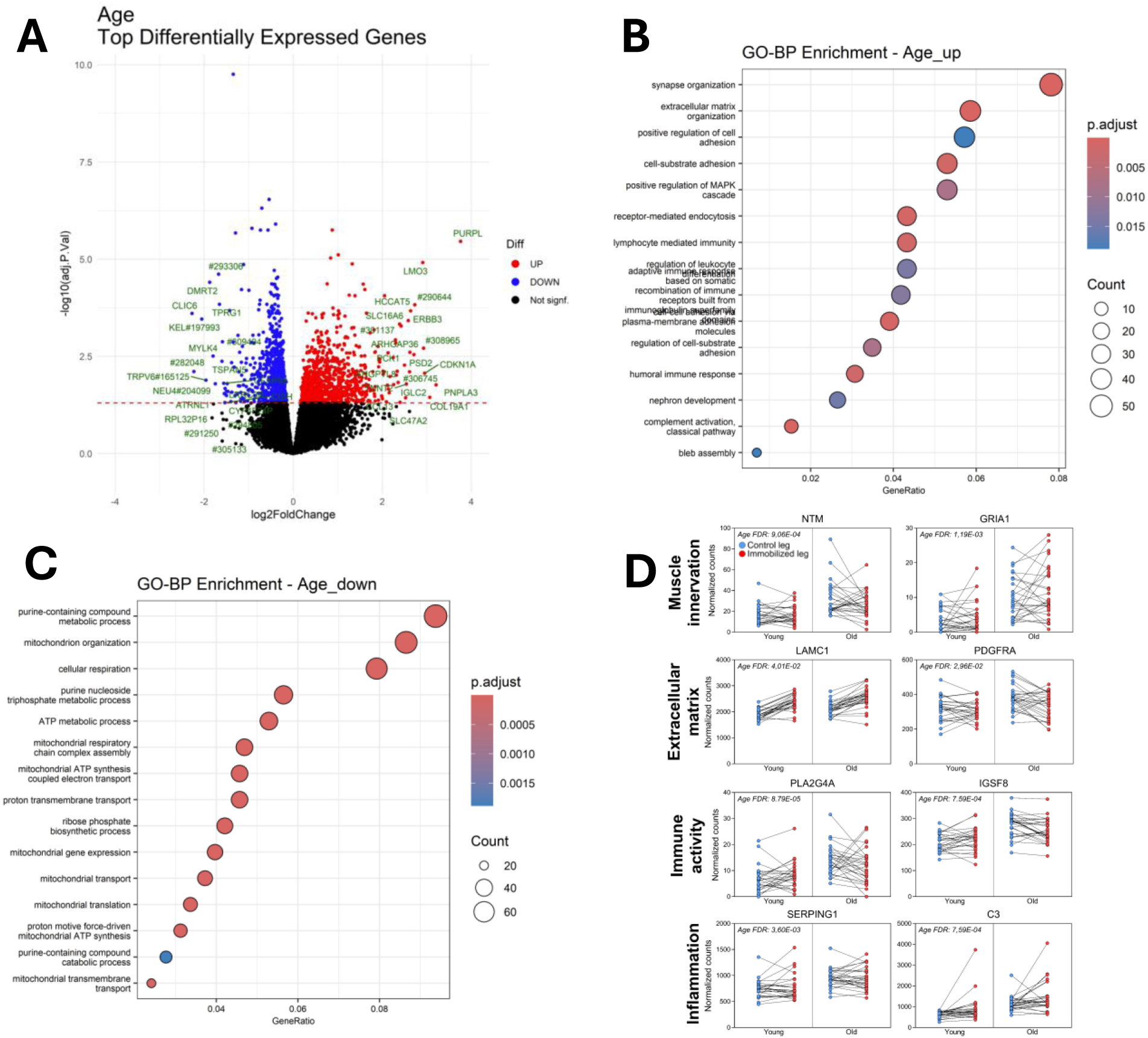
Ageing is associated with neuromuscular, extracellular matrix, immune, and metabolic remodeling in skeletal muscle. (A) Differential expression analysis comparing younger and older participants identified 1954 age-associated genes (FDR<0.05). (B–C) Gene Ontology biological-process enrichment analyses showing pathways upregulated (B) and downregulated (C) with age. (D) Expression of selected genes related to neuromuscular remodeling, extracellular matrix organization, immune activity, inflammation, and mitochondrial metabolism. Differential expression analyses were corrected for multiple testing using the false discovery rate, with significance defined as FDR<0.05. Results are shown with both legs combined; analyses restricted to the control leg yielded comparable findings (Supplementary material 8E).

Pathways upregulated with immobilization were dominated by processes related to proteolysis and cellular stress, including autophagy and ubiquitin-dependent protein turnover, with increased expression of genes such as VCP and FBXO32, consistent with established catabolic and proteostatic responses to human muscle disuse (*34, 35*) (Fig. 3D-E; Data sheet 3). In agreement with the RT-qPCR findings and previous disuse studies (*23, 25*), genes associated with neuromuscular remodeling and denervation-related responses, including CHRND and RUNX1, were also increased (Fig. 3F). Beyond these established responses, the large cohort allowed us to identify pronounced induction of several less well-characterized components of the human immobilization response. Among the most strongly induced genes, MYC increased by approximately 6-fold in the immobilized leg. Given recent evidence that pulsatile MYC expression can promote skeletal muscle hypertrophy (*36*), this increase may reflect a broader compensatory response rather than a purely catabolic signal. CDKN1A, a regulator of cell-cycle arrest and cellular stress responses, and GADD45A, a stress-responsive gene involved in growth arrest and DNA-damage signaling, were also markedly increased, by approximately 10-fold and 13-fold, respectively. Genes related to inflammatory and stress signaling, including IL18, were likewise induced. Notably, despite this extensive stress and remodeling response, we found no clear transcriptional evidence of an immobilization-induced fibrotic program, contrasting with previous reports (*37*).

Downregulated pathways were enriched for muscle system processes, oxidative metabolism, and mitochondrial function (Fig. 3E), consistent with established metabolic deconditioning during human muscle disuse (*38*). Several metabolically relevant genes were reduced, including ETNPPL and SLC16A9, consistent with impaired substrate metabolism during immobilization (*39*). This interpretation was further supported by decreased expression of mitochondrial gene sets (Supplementary Material 8C). Several genes related to muscle structure, remodeling, and contractile function were also downregulated, in agreement with prior disuse studies (*40*). These included sarcomeric and myofibrillar genes such as MYH7, MYLK2, and TTN; membrane-associated structural genes such as DES and CAV3; excitation–contraction coupling genes such as ATP2A1, ATP2A2, and CASQ2; and signaling-related genes such as VEGFB, NOS1, and SMAD7. Many of these genes were represented within Gene Ontology biological-process categories such as “heart contraction” and “striated muscle cell differentiation,” reflecting broad suppression of genes involved in striated muscle structure and function during immobilization. Importantly, these immobilization-associated transcriptional effects were preserved when the analysis was restricted to the No-train group, indicating that the overall disuse response was not driven by inclusion of the Prehab group (Supplementary Material 8D).

Collectively, these findings indicate that short-term immobilization induces a coordinated shift toward a more catabolic, structurally compromised, and metabolically less active skeletal muscle phenotype.

To fully leverage the size of the cohort, including biopsies obtained from both legs of each participant, we also examined the main effect of Age in the RNA-seq dataset. This analysis identified a robust ageing-associated transcriptional signature comprising 1954 DEGs (Fig. 4A). Older muscle showed higher expression of pathways related to neuromuscular remodeling (e.g., GRIA1, NTM), extracellular matrix organization (e.g., PDGFRA, LAMC1), immune activity (e.g., IGSF8, PLA2G4A), and inflammatory signaling (e.g., C3, SERPING1). By contrast, downregulated pathways were predominantly metabolic (e.g., ETFRF1, COX7C, CYCS, SDHC), including oxidative phosphorylation and mitochondrial energy metabolism (Fig. 4B-D). The increased inflammatory and extracellular matrix signatures and reduced mitochondrial metabolism are consistent with established features of ageing human skeletal muscle (*41*). The enrichment of neuromuscular-remodeling pathways further supports emerging evidence that altered neural input and synaptic organization form part of the ageing muscle transcriptome (*42*).

These findings were largely preserved when the analysis was restricted to control-leg biopsies, thereby excluding the immobilized legs (Supplementary Material 8E). This indicates that the ageing-associated signature was not driven by immobilization itself. Moreover, no genes showed significant Age × Leg interactions, suggesting that immobilization elicited broadly similar transcriptional responses in younger and older participants.

Collectively, these findings demonstrate that short-term immobilization elicited extensive transcriptional remodeling in human skeletal muscle despite the physiological benefits of prehabilitation, including activation of catabolic, inflammatory, stress-related, and denervation-associated programs together with suppression of mitochondrial and muscle-structural pathways. Ageing was associated with a distinct but similarly broad transcriptional signature, whereas the molecular response to immobilization was conserved across age groups. Thus, prehabilitation did not prevent the adverse molecular response to disuse, reinforcing the conclusion that its benefits arose primarily from the greater muscle and strength reserve established before immobilization.

### Network analysis identifies biological programs associated with disuse-induced decline

Having established that immobilization induced extensive transcriptional remodeling that was not broadly attenuated by prehabilitation, we next used weighted gene co-expression network analysis (WGCNA) to determine whether these coordinated molecular responses were linked to the accompanying losses in muscle size, morphology, and function (*43*). WGCNA identifies modules of co-expressed genes that can be related to physiological outcomes. Module eigengenes were therefore correlated with *in vivo* measures of muscle function and biopsy-derived indices of muscle morphology to determine which biological programs tracked with disuse-related phenotypic decline.

Sixteen co-expression modules were identified and labeled using conventional color nomenclature (Fig. 5A; Data sheet 4-5). Module–trait analysis showed that several modules were associated with Age, Leg, or both, whereas no module showed a robust association with Group (Supplementary Material 8F). Thus, consistent with the absence of DEGs attributable to prehabilitation, there was no evidence of a coordinated transcriptional program linked to prehabilitation. Several modules were associated with both Age and Leg, and the direction of these associations differed in some cases, indicating that ageing and immobilization may influence shared co-expression programs in distinct ways.

**Figure 5.**
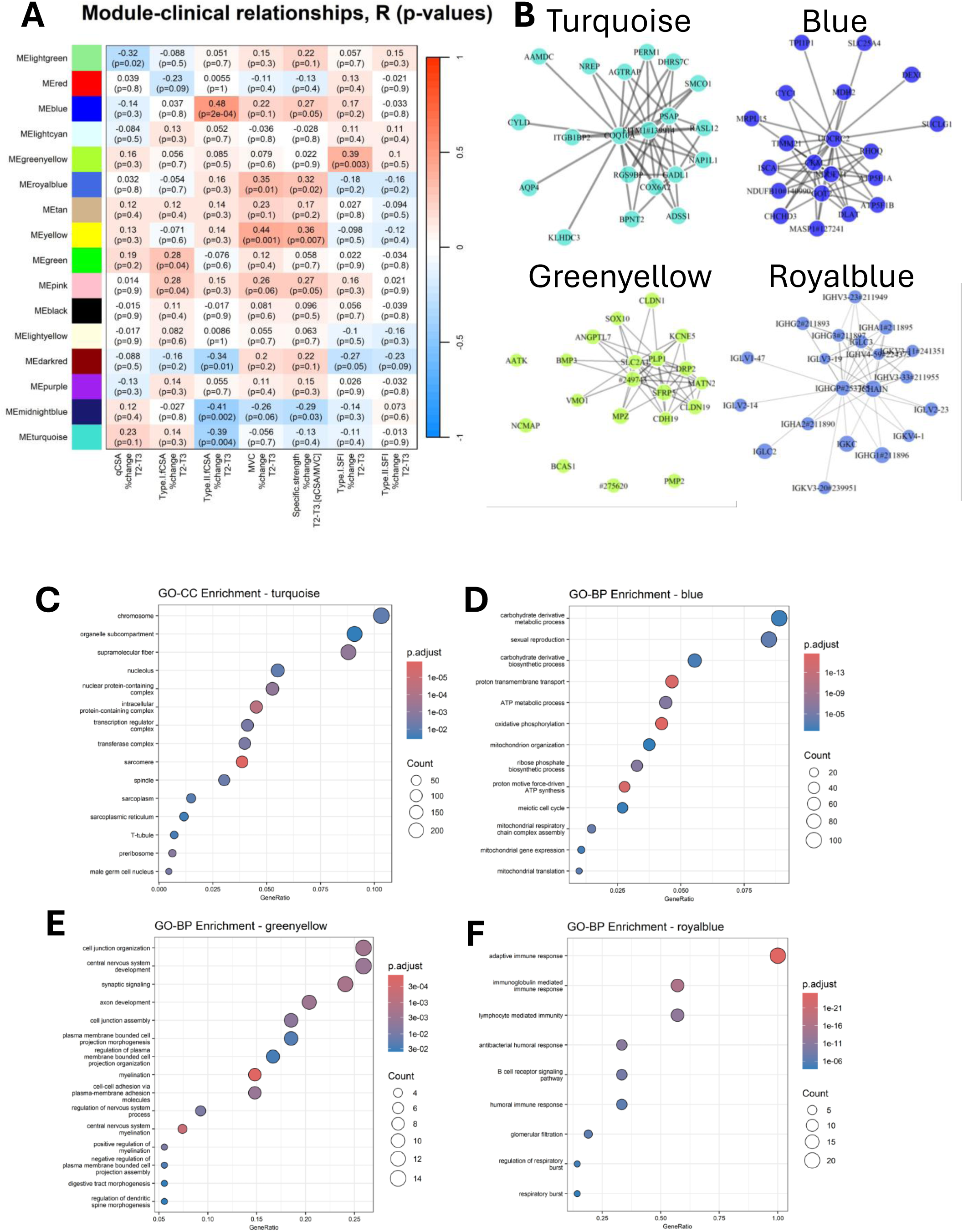
Co-expression network analysis links coordinated disuse-responsive gene programs to changes in muscle morphology and function. (A) Correlations between module eigengenes and immobilization-induced percentage changes in physiological variables from pre- to post-immobilization (ΔT2→T3). Colors indicate Pearson correlation coefficients, with corresponding r and p values shown within each cell. (B) Network visualization of four (out of 16) selected co-expression modules. (C) Turquoise module, enriched for genes related to muscle structure and remodeling. (D) Blue module, enriched for mitochondrial bioenergetics and oxidative metabolism. (E) Greenyellow module, enriched for neuromuscular and structural remodeling. (F) Royalblue module, enriched for immune and inflammatory signaling.

To align the transcriptional and phenotypic measures, percentage changes in physiological outcomes from pre- to post-immobilization were correlated with post-immobilization between-leg differences in module eigengene expression. Across the modules, four broad biological themes emerged; mitochondrial bioenergetics, neuromuscular signaling, immune and inflammatory activity, and structural muscle remodeling (Fig. 5B).

The turquoise module was negatively associated with changes in type II fCSA and was enriched for genes related to structural muscle organization and remodeling (Fig. 5C). The blue module was strongly enriched for genes involved in oxidative phosphorylation and mitochondrial energy production and was positively associated with changes in type II fCSA and specific strength, defined as MVC normalized to quadriceps CSA (Fig. 5D). The greenyellow module was enriched for neuronal and synaptic genes and was associated with changes in type I SFI (Fig. 5E). The royalblue module was enriched for immune-related transcripts and showed associations with changes in MVC and specific strength (Fig. 5F).

The number of significant phenotype associations varied across modules and outcomes. Quadriceps CSA was associated with only one module (lightgreen), whereas specific strength was associated with five modules (blue, royalblue, yellow, pink and midnightblue). Several modules showed no significant phenotype associations, while the pink module showed the greatest number, with associations with type II fCSA, MVC, and specific strength. However, the pink module showed no significant enrichment for Gene Ontology biological process, cellular component, or molecular function terms, indicating that its constituent genes could not be assigned to a single clearly defined biological program.

Together, these findings show that the extensive transcriptional response to immobilization was organized into coordinated programs involving mitochondrial metabolism, immune signaling, neuromuscular processes, and structural muscle remodeling, several of which were associated with concurrent declines in muscle size, strength, and myofiber morphology. The absence of robust module-level associations with prehabilitation further indicates that prehabilitation did not induce a detectable coordinated transcriptional program after immobilization. Because these immobilization-associated molecular programs remained evident despite prehabilitation, the network analysis further supports the conclusion that prehabilitation improved post-disuse outcomes primarily by increasing pre-immobilization reserve, rather than by broadly attenuating the biological response to immobilization.

## Discussion

This preregistered randomized controlled trial provides three main findings. First, older women retained the capacity to respond to short-term high-intensity resistance exercise and did not show evidence of impaired recovery following immobilization. Second, prehabilitation was effective in both younger and older women, primarily by increasing muscle mass and strength before immobilization and thereby establishing greater pre-disuse reserve, rather than by attenuating the relative response to disuse. Third, despite these physiological benefits of prehabilitation, immobilization still induced extensive transcriptional remodeling involving metabolic, immune, neuromuscular, and structural programs, several of which were associated with concurrent changes in muscle size, morphology, and strength. Together, these findings indicate that short-term prehabilitation increases muscle and strength reserve before disuse, thereby preserving a higher absolute level of muscle mass and function after immobilization, while leaving the underlying biological response to immobilization largely intact.

Previous studies have reported delayed or incomplete recovery following disuse in older adults and animal models (5–11), potentially reflecting impaired anabolic and/or neuromuscular responsiveness. In the present study, however, older women responded to prehabilitation and rehabilitation without evidence of impairment relative to younger women. Notably, older participants in the No-train group also returned to baseline following rehabilitation, indicating preserved recovery capacity even without the additional reserve established by prehabilitation. This pattern was broadly consistent across MRI-derived quadriceps CSA, muscle strength, and muscle fiber outcomes. The discrepancy with parts of the existing literature may reflect the healthy and physically active characteristics of the present cohort, including the absence of major chronic disease, relatively high habitual physical activity, and adequate protein intake. These findings suggest that, among otherwise healthy older women able to exercise, chronological age alone does not necessarily limit the capacity to build reserve before disuse or recover following immobilization.

Prehabilitative exercise is receiving increasing scientific and clinical attention (*14–20*), but heterogeneity in study design, patient populations, and assessment timing has made it difficult to determine whether its benefits arise before, during, or after disuse. By assessing participants at baseline, immediately before immobilization, after immobilization, and following rehabilitation, the present study distinguished between these possibilities. Prehabilitation increased muscle size and strength before immobilization, while the relative losses during disuse were similar between groups. Its principal effect was therefore to shift the physiological trajectory upward before immobilization rather than reduce the magnitude of the subsequent decline. The neuromuscular and transcriptomic findings reinforced this interpretation. Immobilization induced marked increases in denervation-associated AChR transcripts (*23, 25, 44*), but prehabilitation did not preserve markers of neuromuscular innervation. Likewise, despite extensive transcriptional effects of immobilization, no genes differed in expression between the Prehab and No-train groups after immobilization. Thus, the physiological benefit of prehabilitation was not accompanied by detectable preservation of the assessed neuromuscular markers or by broad attenuation of the post-immobilization transcriptomic response. Clinically, this supports prehabilitation as a strategy to increase muscle and strength reserve before predictable periods of disuse, such as planned surgery.

Human transcriptomic studies of muscle disuse remain limited (*45*), and have generally relied on pre–post comparisons within the same limb (*23, 25, 34, 35, 38, 40, 46, 47*). By sampling control and immobilized legs at the same post-immobilization time point, the present study enabled direct within-participant comparisons while minimizing temporal and environmental confounding (*48*). Immobilization affected approximately 41% of genes included in the analysis and induced established signatures of proteolysis, mitochondrial suppression, inflammation, denervation, and structural remodeling (*23, 40*). Network analysis further showed that these coordinated programs were associated with concurrent changes in muscle size, morphology, and strength. Less well-characterized features included marked induction of MYC, CDKN1A, and GADD45A and the absence of a clear fibrotic transcriptional program (*36, 37, 49, 50*). Collectively, these findings demonstrate that extensive adverse molecular remodeling remained evident despite prehabilitation, further supporting reserve accumulation rather than biological resistance as the principal mechanism of benefit.

Aging was associated with increased neuromuscular, extracellular matrix, immune, and inflammatory signaling together with reduced oxidative and mitochondrial metabolism, broadly consistent with previous transcriptomic studies of ageing muscle (*51*). However, no genes showed significant Age × Leg interactions, and the physiological response to immobilization was also broadly comparable between age groups. This contrasts with Mahmassani et al., who reported age-dependent transcriptional responses to 5 days of bed rest, including greater dysregulation of fibrosis, inflammation and mechanosensing pathways in older adults (*34*). Thus, despite marked baseline differences in the muscle transcriptome, younger and older women exhibited broadly similar molecular and physiological responses to short-term immobilization.

This study has several limitations. First, the study was conducted in healthy volunteers rather than surgical or clinical populations. Although this controlled experimental model was necessary to isolate the relative contributions of reserve accumulation, disuse-induced decline, and subsequent recovery, it may limit direct generalizability to patients exposed to additional perioperative or disease-related stressors. Second, quadriceps CSA and muscle biopsies were assessed at different points during immobilization, with MRI performed after 5 days and biopsies collected after 8 days for logistical reasons. The physiological and molecular measurements therefore represent closely related, but not identical, stages of disuse. Third, primary cell cultures were derived exclusively from older participants, precluding direct age-group comparisons at the cellular level. In addition, the cohort consisted solely of women, which improved internal consistency but may limit generalizability to men. Although menstrual regularity and hormonal contraceptive use were recorded at screening, menstrual cycle phase was not standardized or incorporated into the analyses, which may have introduced additional biological variability. Finally, ten participants withdrew before study completion, representing a relatively high attrition rate.

In conclusion, short-term prehabilitation improved outcomes after limb immobilization in younger and older women primarily by increasing muscle and strength reserve before disuse. It did not materially attenuate the relative physiological decline, preserve neuromuscular innervation, or alter the broad molecular response to immobilization. These findings suggest that the principal value of prehabilitation lies in raising the functional starting point before predictable disuse rather than rendering skeletal muscle resistant to the disuse process itself. They therefore support prehabilitation as a potential strategy to increase muscle and strength reserve before planned periods of immobilization.

## Materials and Methods

### Study design

This was a single-center, parallel-group, randomized, controlled, outcome-assessor-blinded trial conducted at the Institute of Sports Medicine Copenhagen, Copenhagen University Hospital - Bispebjerg and Frederiksberg, Copenhagen, Denmark, in accordance with CONSORT guidelines (*52*). Participants were assigned to either four weeks of supervised heavy resistance training (Prehab) or to maintain their habitual lifestyle (No-Train). This was followed by 8 days of unilateral lower-limb immobilization with a knee brace. Subsequently, all participants undertook a four-week period of heavy resistance training (rehabilitation). The total study duration for each participant was 10 weeks.

Testing was organized into four main time points: baseline, post-prehabilitation/pre-immobilization, post-immobilization, and post-rehabilitation. Each time point consisted of two test days. On the first day, participants underwent MRI scanning, and on the second day they completed a DXA scan (only at baseline), blood sampling, muscle biopsies, and maximal voluntary contraction (MVC) testing. For the post-immobilization time point, MRI was acquired on immobilization day 5 due to scanner availability, whereas blood sampling, muscle biopsies, and MVC were performed on immobilization day 8 (immediately after brace removal). An additional mid-immobilization visit on day 3 included blood sampling only. A familiarization session was conducted 1–2 weeks before baseline testing, during which participants completed MVC testing, hand-grip strength, and the chair-stand test. In total, each participant attended 10 visits in addition to the supervised training sessions.

### Ethics and registration

The trial was approved by the Committees on Health Research Ethics for the Capital Region of Denmark (Ref: H-23040446; approval date: 24 August 2023). All participants provided written informed consent prior to enrollment. The study was preregistered at www.ClinicalTrials.gov (NCT06205784; registration date: 19 December 2023). Amendments to the protocol included lowering the lower age limit of the older group from 65 to 60 years (approved by the ethics committee but not updated in the ClinicalTrials record) and increasing the planned sample size to account for dropouts. The trial was conducted in accordance with the principles of the Declaration of Helsinki.

### Recruitment and enrollment

Participants were recruited through advertisements in local newspapers, on Facebook, and via the department website. Advertisements provided a phone number and email address for direct contact with the principal investigator (C.S.). Following an initial eligibility check, interested individuals were invited to an on-site screening visit, where they could be accompanied by a support person (e.g., partner, friend, or family member). At this visit, full trial details were explained, eligibility was assessed, and participants were then given time to consider their participation. Enrollment was completed only after written informed consent was obtained. Recruitment began in January 2024, and the final participant completed the study in June 2025.

### Eligibility criteria

Eligible participants were community-dwelling women aged 20–35 years or 60–85 years, with a body mass index (BMI) of 18.5-35 kg/m², who were generally healthy and consumed a non-vegan diet. Exclusion criteria were: Current or recent regular smoking; current pregnancy or pregnancy within the previous 3 months; current or past drug or alcohol abuse; a history of one or more muscle biopsies from both vastus lateralis muscles; medical conditions likely to affect protein metabolism or interfere with trial participation; knee pain limiting the ability to train; use of strong blood-thinning medications; regular and high intake of substances with anticoagulant properties (e.g., fish oil, ginger, ibuprofen); regular resistance training within the past year; regular participation in intense exercise at the time of enrollment; prior intake of deuterium oxide; contraindications to MRI due to metal implants; and premenopausal status in participants aged 60–85 years.

### Randomization and blinding

Participants were block-randomized in a 1:1 ratio to prehabilitation (Prehab) or No-Train using a lot-drawing envelope method. Within each participant, the dominant or non-dominant leg was randomly assigned with equal probability to immobilization (the “immobilized leg”); the contralateral leg served as the “control leg”. Leg allocation was concealed until assignment. Blinding was applied where feasible. Image analysts for MRI (primary outcome) and DXA, and laboratory personnel processing biopsy material (histology and RNA sequencing), worked from coded files and were blinded to Age, Group, and Time. In contrast, assessment of performance tests (e.g., muscle strength assessments) was not blinded. Participants and exercise supervisors were unblinded to Group, and immobilization was apparent.

### Intervention and immobilization

#### Prehabilitative exercise

Participants randomized to the Prehab group completed 10 supervised sessions of heavy resistance training (HReT) over a 4-week period. Study personnel maintained a training diary for each participant, documenting machine settings, load, repetitions, and sets for every exercise. From the third session onward, participants were encouraged to train close to volitional failure. Each session began with a 5-min warm-up on a stationary cycle ergometer, followed by three lower-body exercises (leg press, knee extension, leg curl) and two upper-body exercises (pulldown and shoulder press; MED Line, Technogym, Gambettola, Italy). Leg press and knee extension were performed unilaterally, with the starting leg alternating between sessions. For approximately three weeks, the seated leg-curl machine was out of service and was temporarily replaced with a lying leg-curl machine. The preferred exercise sequence was maintained when possible, though minor adjustments were permitted for logistical reasons. Rest periods were timed with stopwatches and were ∼90 s between sets in sessions 1–2 and ∼120 s from session 3 onward. Each training session lasted approximately 45–75 min. One-repetition maximum was assessed for leg press and knee extension before the first and final training sessions. A minimum attendance of 9 of 10 sessions was required for inclusion in the per-protocol analyses. The full training protocol is provided in Supplementary Material 9.

#### Rehabilitative exercise

The rehabilitation program was identical to the prehabilitation protocol and was undertaken by participants in both the Prehab and No-Train groups. During this phase, participants originally assigned to Prehab did not have access to their prehabilitation training logs, except for machine settings needed to ensure correct equipment setup.

#### Immobilization

Lower-limb immobilization was achieved by fixing the knee joint at 75° of flexion (0° = full extension) using a hinged knee brace (#807A, Albrecht GmbH, Bernau am Chiemsee, Germany). The brace was worn continuously for 8 days, including during sleep and bathing, and was secured with sealing strips to prevent removal. The only exception was during the MRI scan on day 5, when the brace was removed for imaging. Participants were transported to and from the scanner by wheelchair and were instructed not to bear weight on the immobilized leg at any time during the procedure. To eliminate weight-bearing on the immobilized leg, participants were provided with height-adjusted crutches and received thorough instruction in their use, including stair navigation; those unable to use crutches safely were offered a four-footed walker. Participants were encouraged to perform hourly “venous-pump” ankle exercises (active dorsiflexion–plantarflexion for approximately 1–2 minutes) and to remain otherwise as active as safely possible (e.g., regular walking with their assistive device). Compression stockings were provided for all participants and were recommended for continuous use throughout the 8-day intervention in older participants. The shoe was removed from the immobilized leg throughout the intervention to minimize inadvertent loading. Participants were also advised to maintain good hydration, particularly during warm weather, and to limit alcohol intake to reduce the risk of dehydration. For travel to and from the study site during the immobilization period, taxi transport was arranged to minimize unnecessary physical activity, and wheelchairs were available at the study site for participants who preferred them. Adherence to immobilization was assessed by step-count monitoring over a 4-day period (day 4 to 7 of immobilization) using an accelerometer (activPAL micro, PAL Technologies, Glasgow, Scotland) affixed to the non-immobilized leg. Participants were instructed to promptly report any discomfort, skin irritation, or brace-related problems, and at each follow-up visit study staff inspected the immobilized limb and brace positioning to ensure both safety and tolerability.

### Outcomes

The primary outcome was the change in quadriceps cross-sectional area (CSA), assessed by magnetic resonance imaging, from baseline to post-immobilization and post-rehabilitation. A number of secondary outcomes were prespecified in the trial registry. The present study focuses on those most directly related to muscle mass and strength, namely muscle fiber size, denervated fibers and muscle gene expression.

#### Muscle mass

MRI was used as the gold-standard method for *in vivo* assessment of muscle mass owing to its high reliability and validity (53). Scans were performed at Department of Radiology, Copenhagen University Hospital - Bispebjerg and Frederiksberg, Copenhagen, Denmark, by a core group of four radiographers, all blinded to group allocation. Participants were scanned in the supine position, and only the immobilized leg was imaged. A 3.0 T MRI (Vida: Siemens Healthineers, Erlangen Germany) scanner was used. A T1-weighted sequence was acquired, consisting of 50 transverse slices with a slice thickness of 5 mm and an inter-slice gap of 0.5 mm (5.5 mm center-to-center). The stack started at the level of the medial-tibial plateau and extended proximally, covering a total length of 27.5 cm. The image analysis approach has been described previously (54). For analysis, slice 40, located 22 cm proximal to the tibial plateau and corresponding approximately to the mid-femur, was selected for all participants, with minor adjustments made based on anatomical landmarks to ensure consistency across time points. Contrast and brightness were standardized before analysis. Using Horos software (Horosproject.org, Nimble Co LLC d/b/a Purview, Annapolis, MD, USA), the borders of vastus lateralis, rectus femoris, vastus medialis, and vastus intermedius were manually delineated, and quadriceps cross-sectional area (CSA) was defined as the sum of these muscles. All scans for each participant were analyzed together by an assessor blinded to Age, Group and Time. Each image was analyzed twice, and the average value was used. If the coefficient of variation (CV) between the two measurements exceeded a predefined threshold (CV >2% for rectus femoris and >1.5% for the other muscles), a third analysis was performed and the most discrepant value was discarded.

#### Muscle strength

Maximal voluntary contraction (MVC) of the knee extensors in the immobilized leg was assessed at all four time points (baseline, pre-immobilization, post-immobilization, and post-rehabilitation). Testing was performed on a Kinetic Communicator dynamometer (KinCom, model 500-11, Chattecx, Chattanooga, TN), as previously described (*28*). Isometric MVCs were obtained at a knee angle of 70° flexion (0° = full extension).

The original protocol included isokinetic MVCs at slow and fast angular velocities; however, a mid-study breakdown of the KinCom necessitated replacement with a Biodex System 4 (Biodex Medical Systems, Shirley, NY, USA). Because the two devices differ in torque sampling, subsequent testing on the Biodex was limited to isometric MVCs to maximize methodological comparability. A post hoc device comparison in independent volunteers (n=8) demonstrated that KinCom and Biodex were not directly comparable (percent-difference Bland–Altman mean bias ∼+10.9%, with wide limits of agreement), and thus primary strength analyses were restricted to participants tested on the same device across all time points. Mixed-device data are presented in Supplementary Material 6.

#### Lean body mass

Whole-body dual-energy X-ray absorptiometry (DXA) was used to assess body composition at baseline. All scans were performed on a Lunar DPX-IQ scanner (GE Healthcare, Chalfont St. Giles, UK), as previously described (54).

#### Blood

Venous blood samples were drawn from the antecubital vein at all four main study time points (baseline, pre-immobilization, post-immobilization, and post-intervention), as well as on day 3 of the immobilization period. Samples were analyzed for markers of immune function (leukocytes, thrombocytes), systemic inflammation (C-reactive protein), and fluid homeostasis (potassium, creatinine, sodium). All analyses were performed according to standard procedures at the Department of Clinical Biochemistry, Bispebjerg Hospital.

#### Muscle biopsies

Muscle biopsies were obtained from the mid-portion of the vastus lateralis under local anaesthesia (lidocaine) using a 5- or 6-mm Bergström needle fitted with manual suction (*55*). After a small skin incision, samples were collected in a randomized pattern to avoid local effects of repeated sampling (*56*). In total, five biopsies were collected from each participant, four from the immobilization leg, one at each study time point, and one from the control leg at the Post-immobilization time point. The extracted tissue was divided for histological analyses and, at the post-immobilization time point only, primary cell culture. For histological analyses, intact pieces were gently aligned in Tissue-Tek® (Sakura Finetek), rapidly frozen in isopentane (JT Baker) pre-cooled in liquid nitrogen, and stored at −80 °C until sectioning.

#### Muscle biopsy analyses

For histological analyses, muscle tissue blocks were cryosectioned at 10 µm thickness. Sections from the baseline, pre-immobilization and post samples were mounted on the same glass slide, whereas the two samples obtained post-immobilization (immobilization and control leg) were mounted on a separate slide. Both slides were stained together to minimize batch effects. All immunohistochemical stainings were completed across two staining rounds. Antibody details are provided in Supplementary material 11.

To identify denervated fibers, cryosections were stained with anti-CD56/NCAM (347740; Becton Dickinson) and anti-dystrophin (#D8168; Sigma–Aldrich), as previously described (*57*). Primary antibodies were diluted in TBS containing 1% BSA and incubated overnight at 4°C. Sections were then washed twice for 5 min in TBS and incubated with the appropriate secondary antibodies, diluted in TBS containing 1% BSA, for 45 min at room temperature. Following two additional washes in TBS, sections were fixed in Histofix (Histolab, Sweden) and mounted with ProLong Gold Antifade Mountant containing DAPI. Quantifications were expressed relative to the total number of fibers within each section. On average, 1921 ± 852 fibers per sample (range 394–4,830) were analyzed. For determination of fiber-type composition and morphology, sections were stained with anti-MyHC-I (DSHB, BA-D5) and anti-MyHC-IIa (DSHB, SC-71), together with wheat-germ agglutinin (WGA; Invitrogen, W32465) to delineate cell borders. The staining procedure followed the same general protocol as described above, except that secondary antibodies were incubated for 60 min. These stainings were used to quantify fiber-type distribution, cross-sectional area, and fiber shape metrics as previously described (*58*). On average, 479 ± 92 fibers per sample (range 178–745) were analyzed.

For both NCAM and MyHC immunofluorescence, whole-section imaging was performed using an AxioScan.Z1 slide scanner (Carl Zeiss). A region of interest was manually defined around the tissue, and the entire section was acquired using sequential single-channel imaging with coarse and fine autofocus steps.

Tiles were captured with 10% overlap and subsequently stitched using ZEN Blue software (Carl Zeiss). Image analyses were conducted in Fiji (*59*), using the ObjectJ plugin for identification of denervated fibers and custom semi-automated macros for fiber morphology and shape analyses as previously described (*60*).

#### qPCR

Approximately 100 cryo sections of 10 μm thickness (around 5-10 mg) from the embedded tissue were homogenized in 1 mL of TriReagent (Molecular Research Center, Cincinnati, OH, USA) containing five 2.3 mm stainless steel balls and one 1.0 mm Silicon Carbide bead (BioSpec Products, Bartlesville, Oklahoma, USA) by shaking in a FastPrep®-24 instrument (MP Biomedicals, Illkirch, France) at speed level 4 for 15 s. Following homogenization, bromo-chloropropane was added in order to separate the samples into an aqueous and an organic phase. Following isolation of the aqueous phase, RNA was precipitated using isopropanol. The RNA pellet was then washed in ethanol and subsequently dissolved in 20 μL RNAse-free water. Total RNA concentration and purity was determined using a DS-11 FX+ spectrophotometer (DeNovix, Wilmington, DE, USA).

350 ng total RNA was converted into cDNA in 20 μL using the qScript cDNA Supermix (Quantabio, Beverly, MA, USA) and 1 μM poly-dT (Invitrogen, Naerum, Denmark) according to the manufacturer’s protocol (Quantabio). For each target mRNA, 0.25 μL cDNA was amplified in a 25 μL SYBR Green polymerase chain reaction (PCR) containing 1 × Quantitect SYBR Green Master Mix (Qiagen) and 100 nM of each primer (Supplementary Material 10). The amplification was monitored real time using the CFX96 Real-time PCR machine (Bio-Rad, Hercules, CA, USA), protocol 10’, 95° {15’’, 95°; 30’’, 58°; 90’’, 63° (signal collection)} x 50, Melt curve. The Ct values were related to a standard curve made with known concentrations of DNA oligonucleotides (Ultramer oligos, Integrated DNA Technologies, Leuven, Belgium) with a DNA sequence corresponding to the sequence of the expected PCR product. The specificity of the PCR products was confirmed by melting curve analysis after amplification. RPLP0 mRNA was chosen as internal control. Data are shown relative to the geometric mean of the Pre samples from all subjects.

#### RNA sequencing and bioinformatics

RNAseq was performed by a commercial company (Azenta, Liepzig, Germany). Briefly, RNA samples were quantified using Qubit 4.0 Fluorometer (Life Technologies, Carlsbad, CA, USA) and RNA integrity was checked with RNA Kit on Agilent 5300 Fragment Analyzer (Agilent Technologies, Palo Alto, CA, USA).

ERCC RNA Spike-In Mix from ThermoFisher Scientific, was added to normalized total RNA prior to library preparation following manufacturer’s protocol. RNA sequencing library preparation was prepared using NEBNext Ultra II Directional RNA Library Prep Kit for Illumina following manufacturer’s instructions (NEB, Ipswich, MA, USA). Briefly, mRNAs were first enriched with Oligo(dT) beads. Enriched mRNAs were fragmented. First strand and second strand cDNA were subsequently synthesized. The second strand of cDNA was marked by incorporating dUTP during the synthesis. cDNA fragments were adenylated at 3’ends, and indexed adapter was ligated to cDNA fragments. Limited cycle PCR was used for library amplification. The dUTP incorporated into the cDNA of the second strand enabled its specific degradation to maintain strand specificity. Sequencing libraries were validated using NGS Kit on the Agilent 5300 Fragment Analyzer (Agilent Technologies, Palo Alto, CA, USA), and quantified by using Qubit 4.0 Fluorometer (Invitrogen, Carlsbad, CA). The sequencing libraries were multiplexed and loaded on the flow cell on the Illumina NovaSeq X plus instrument according to manufacturer’s instructions. The samples were sequenced using a 2x150 Pair-End (PE) configuration v1.5. Image analysis and base calling were conducted by the NovaSeq Control Software v1.7 on the NovaSeq instrument. Raw sequence data (.bcl files) generated from Illumina NovaSeq was converted into fastq files and de-multiplexed using Illumina bcl2fastq program version 2.20. One mismatch was allowed for index sequence identification.

The sequencing quality was confirmed with FastQC v0.11.9 and reads were cleaned and deduplicated using Fastp v0.23.4 (*61*). They were then aligned to the Human GRCh38.p14 genome and transcripts (Gencode v47, exons only) counted using SubRead v2.0.3 (*62*) resulting in 14–24 million counts per sample. 40054 transcripts were identified after filtering for at least ten total counts. The RNAseq data have been deposited in Array-Express with the data set identifier E-MTAB-17648. Mixed effects model, Group x Age x Leg + (1 | Subject), was used to find differentially expressed genes using the Dream procedure (*63*) from variancePartition v1.38.1 (parameters: min.count = 10, min.total.count = 15, large.n = 10, min.prop = 0.7). WGCNA analysis was performed using WGCNA v1.74 (*43*). Parameters; RsquareCut = 0.8, minClusterSize = 20, cutHeight = 0.25). Counts were vst normalized (transcripts with mean < 1 or var < 0.1 excluded) before the WGCNA analysis. GO-term enrichment analysis was performed using topGO v2.60.1, with the elim algorithm and fisher test (*64*). Only genes assigned an actual FDR value were included in the pathway analysis (the background list).

#### Cell culture

For *in vitro* experiments, primary mononucleated cells were isolated from fresh muscle biopsy tissue using a modified version of a published protocol (*65*). Briefly, dissected tissue was enzymatically dissociated for 1 h in a continuously agitated digestion buffer containing collagenase II and dispase II, after which the liberated cells were collected and plated in T-25 or T-75 culture flasks. Cells were expanded in growth medium (C-23060; PromoCell) supplemented with 1% L-glutamine–penicillin–streptomycin solution (GPS; G6784; Sigma) and 15% fetal bovine serum (FBS; ALB-S1810; Biowest) for 7–10 days, with medium changes every 2–3 days. At the end of this initial expansion period, cells were detached from the flasks, cryopreserved using controlled-rate “Frosty-Boy” freezing containers at –80°C, and transferred to long-term liquid-nitrogen storage the following day.

Primary cell-culture experiments were performed using samples from older participants only. This decision was made to focus resources on the cohort in which age-related impairments in muscle regenerative capacity were expected. Cells were stored in liquid nitrogen for approximately 6–12 months before experimentation. Cryopreserved cells were rapidly thawed, plated in T-75 flasks in growth medium, and expanded for 3 days. Cells were then sorted using CD56 magnetic microbeads (130–050-401; Miltenyi Biotec), as previously described (*31*), yielding a CD56 fraction enriched in muscle stem cells (MuSCs) and a CD56 fraction consisting predominantly of fibroblasts.

Sorted MuSCs were subsequently used in three downstream assays. For the proliferation assay, MuSCs were seeded in growth medium at a density of 3000 cells/cm² on glass coverslips in 24-well plates and cultured for 3 days. Cells were then fixed with Histofix for 10 min, and immunostained for Desmin (Abcam, ab32362) to identify myogenic cells and Ki67 (Abcam, ab238020) to assess proliferative activity. Cultures were imaged using a DP71 camera (Olympus) mounted on a BX51 microscope (Olympus) with a 10× objective. Four images were acquired per sample from predefined positions north, south, east, and west of the coverslip center. A mean of 1109 ± 154 cells were analyzed per sample (range 530–1362). The proliferation index was calculated as the proportion of Desmin-positive cells co-expressing Ki67 relative to the total number of Desmin-positive cells.

For the fusion assay, MuSCs were seeded at 25000 cells/cm² on glass coverslips in 24-well plates. The following day, growth medium was replaced with differentiation medium (C-23260; PromoCell) supplemented with 1% GPS, and cells were cultured for a further 4 days, with one medium change midway through the differentiation period. Cells were fixed with Histofix for 10 min and stained for Desmin and Myogenin (DSHB, F5D) to assess myogenic differentiation and myotube formation. Images were acquired using the same microscopy setup and sampling pattern as for the proliferation assay, with four predefined fields obtained per sample. A mean of 297 ± 137 cells were analyzed per sample (range 65–568). The fusion index was calculated as the proportion of nuclei contained within Desmin-positive myotubes comprising ≥3 nuclei relative to the total number of nuclei within Desmin-positive cells.

For the MuSC–motor neuron co-culture assay, sorted MuSCs were seeded and differentiated using the same conditions as for the fusion assay. Primary motor neurons were isolated from embryonic day 15 Sprague–Dawley rat embryos as previously described (*31*). Briefly, spinal cords were dissected, mechanically and enzymatically dissociated, and subjected to density-gradient centrifugation to enrich for motor neurons. A total of six independent motor neuron isolations were performed, each representing a separate experimental batch. Within each batch, MuSCs from both Prehab and No-Train participants and from both the immobilized and control legs were included. Motor neurons were plated directly onto the differentiated MuSC cultures at 5000 cells/cm². At the time of motor neuron addition, 50% of the differentiation medium was replaced with Neurobasal medium, and the co-cultures were maintained for 24 h. Cultures were fixed with Histofix for 10 min and stained for choline acetyltransferase (ChAT;Millipore, AB144P) and Tau-1 (GeneTex, GTX130462) to identify motor neurons. Although both ChAT and Tau-1 were included in the staining protocol, motor neuron quantification was based on Tau-1 because ChAT staining was not sufficiently distinct for reliable quantification in the co-cultures. Cells were imaged using the same microscopy setup and four-field sampling pattern as for the proliferation and fusion assays. Motor neuron abundance was quantified as the number of Tau-1-positive neurons per imaged area. Imaging and subsequent analyses were performed blinded to Leg and Group, and quantitative assessments were carried out using the ObjectJ plugin for Fiji (*59*). Because substantial between-experiment variation was observed across the six independent motor neuron preparations, motor neuron abundance was adjusted for experimental batch by subtracting the mean value of the corresponding batch from each individual measurement and adding the global mean across all six experiments before statistical analysis.

#### Diet registration

To screen for inadequate protein intake, participants conducted diet registrations over four consecutive days during the 1–2 weeks preceding study commencement (two weekdays and two weekend days) using commercial software (www.madlog.dk). 16 participants (5 younger and 11 older) consumed <1.0 g/kg, and were asked to increase daily protein intake, and given recommendations for how to do this.

#### Activity monitoring

The total number of steps performed, and the time spent standing and walking, was measured over four consecutive days (three weekdays and one weekend day) using an accelerometer (activPal micro, PAL technologies, Glasgow, Scotland), that was placed on the thigh of the control leg. Measurements were made during the 1–2 weeks preceding study commencement and during the immobilization (day 4 to 7 of immobilization).

#### Physical testing

A 30-s chair-stand test was performed at baseline to assess lower-body functional strength. Participants stood from a standard-height chair (same model for all tests) with feet shoulder-width apart and arms crossed over the chest. After familiarization, participants completed a single recorded trial, and the number of full stands completed in 30 s was counted.

Maximal hand-grip strength was assessed using a Baseline BIMS Digital Grip Dynamometer (Fabrication Enterprises Inc., White Plains, NY, USA). Participants were seated in a chair with the test arm flexed to approximately 90° at the elbow and the forearm pointing directly forward, without external support. After several practice squeezes to ensure familiarity with the device, participants performed three maximal voluntary contractions with the dominant hand, separated by brief rest periods. The average of the three trials was used for analysis.

#### Questionnaires

At baseline, participants were screened for sarcopenia using the SARC-F questionnaire (66), and their habitual physical activity levels were assessed using the Global Physical Activity Questionnaire (GPAQ) (67).

#### Adverse events

Adverse events were monitored throughout the study. At each visit, participants were specifically asked about new symptoms, discomfort, or health changes, and immobilization-related issues (e.g., skin irritation, brace fit) were checked. In addition, each participant received a phone call during the immobilization period to inquire about their well-being and any problems with the intervention. All reported events were documented and classified as serious or non-serious according to predefined criteria.

### Sample size and statistical analyses

Sample size was determined based on the primary outcome of quadriceps CSA, assessed from baseline to post-immobilization and post-rehabilitation. Based on previous immobilization and resistance-training studies (4, 54, 68), we anticipated an average reduction in CSA of ∼3% in the No-train group following short-term immobilization and no reduction in the Prehab group. To detect a between-group difference of 3 percentage points in CSA change, assuming an SD of 3 percentage points, 80% power, and a two-sided α level of 0.05, 12 participants per group were required. To account for potential attrition, we aimed to enroll 14–16 participants per group, corresponding to 28–32 participants within each age group.

All analyses were conducted according to the per-protocol principle and included participants who completed the intervention and outcome assessments as planned. Data distributions and potential outliers were evaluated by visual inspection. The final sample size is reported for each analysis.

Baseline characteristics were analyzed using the observed values. Normally distributed continuous variables were analyzed using two-way ANOVA with Age Group (younger vs older), Group (Prehab vs No-train), and their interaction as fixed factors. Categorical variables were analyzed using Fisher’s exact test on collapsed 2 × 2 contingency tables. Continuous variables with non-normal distributions were analyzed using Mann– Whitney U tests after collapsing across the other factor to evaluate the main effects of Age or Group.

For analyses of intervention effects, outcomes were expressed as percentage change from baseline, except for fiber-type distribution, NCAM-positive fibers, RT-qPCR and cell-culture endpoints, which were analyzed using the measured values. RT-qPCR data were log-transformed prior to statistical analysis. The primary analytical approach was a three-factor linear mixed-effects model with Time (repeated factor; baseline, pre-immobilization, post-immobilization, and post-rehabilitation), Group (Prehab vs No-train), and Age (younger vs older) as fixed factors, including all two-way interactions and the Time x Group x Age interaction. Participant was included as a random effect. When a significant Group × Time interaction was detected, follow-up comparisons between the Prehab and No-train groups were performed at the pre-immobilization, post-immobilization, and post-rehabilitation time points using unpaired t tests.

The proportion of NCAM-positive fibers showed a non-normal distribution and was therefore analyzed using non-parametric methods. Changes over time were evaluated separately within each group using the Friedman test. Baseline differences between younger and older participants were assessed using the Mann–Whitney U test.

Continuous data are generally presented as mean ± SD unless otherwise stated. Non-parametric data are presented as median with 95% confidence intervals or range. Statistical significance was set at p<0.05. All analyses were performed using GraphPad Prism (GraphPad Software, San Diego, CA, USA).

## Acknowledgments

The monoclonal antibodies BA.D5, and SC-71, both developed by Schiaffino S., were obtained from the Developmental Studies Hybridoma Bank, created by the Eunice Kennedy Shriver National Institute of Child Health and Human Development of the NIH and maintained at The University of Iowa, Department of Biology, Iowa City, IA, USA. We acknowledge the Core Facility for Integrated Microscopy, Faculty of Health and Medical Sciences, University of Copenhagen, for assistance with slide-scanner imaging. We thank the Department of Clinical Biochemistry, Bispebjerg Hospital, University of Copenhagen, for blood analyses, and Jens Hannibal, and the animal-care staff for support with animal housing and handling. We further acknowledge the Department of Radiology, Copenhagen University Hospital – Bispebjerg and Frederiksberg, Copenhagen, Denmark, for MRI acquisition. We thank the Department of Occupational Therapy and Physiotherapy, Bispebjerg Hospital, Copenhagen, Denmark, for providing training facilities.

We are grateful to Anja Jokipii-Utzon and Ann-Christina Ronnié Reimann for excellent technical assistance with muscle biopsy processing for histology and RNA sequencing. Finally, we sincerely thank all students and study participants for their time, commitment, and enthusiasm, without whom this study would not have been possible.

## Data availability

All study data will be made publicly available upon publication. Tabular data will be provided as comma-separated values (CSV) in anonymized, de-identified form, and sequencing datasets will be deposited in the EMBL-EBI ArrayExpress repository (https://www.ebi.ac.uk/biostudies/arrayexpress) under accession number E-MTAB-17648

## Author contributions

C.S., S.P.M., and A.L.M. conceived and designed the research.

C.S., S.D.P., F.H.L., A.E.K., H.L.C., and L.P. performed experiments.

C.S., H.L.C., A.E.K., R.B.S., P.S., and L.P. analyzed data.

C.S., M.K., S.P.M., R.B.S., P.S., and A.L.M. interpreted results of experiments.

C.S. and P.S. prepared figures.

C.S. drafted the manuscript.

All authors edited, revised, and approved the final version of the manuscript.

## Supplementary material

Supplementary material 1: Study flow diagram

Flow of participants through screening, allocation to the Prehab or No-train groups, immobilization, follow-up, and inclusion in the final analyses.

Supplementary material 2: Baseline muscle characteristics by age group

Data are presented as mean±SD. ¹ Young n=17; Older n=20. Abbreviations: CSA, cross-sectional area; fCSA, muscle fiber cross-sectional area; MVC, maximal voluntary contraction; SFI, shape factor index.

Supplementary material 3: Physical activity during immobilization

Daily step count (A) and time spent walking (B) during immobilization, expressed relative to the pre-immobilization period (%). Data are shown as mean ± SD with individual values.

Supplementary material 4: Routine Blood Biomarkers

Venous blood samples were obtained from the antecubital vein at baseline (Pre), pre-immobilization (Pre-Immb), day 3 of immobilization (Peri-Immb), post-immobilization (Post-Immb), and post-recovery (Post). Samples were analyzed for markers of potassium (A), creatinine (B), sodium (C) and C-reactive protein (D; CRP). Individual participant are shown with connected data points across time. Horizontal dotted lines indicate upper and lower reference values. CRP is displayed on a log10 scale; values below the assay lower limit of quantification (1 mg/L) are plotted at 0.5 mg/L.

Supplementary material 5: Adverse events, incidental findings, and safety monitoring Supplementary material 6: Maximal voluntary contraction analyses including all participants

(A) Maximal voluntary contraction (MVC) of the knee extensors measured isometrically at baseline, pre-immobilization, post-immobilization, and post-rehabilitation using measurements obtained from both dynamometers (n=54). Data are shown as mean ± SD. Statistical analyses were performed using linear mixed-effects models with Time, Age, and Group as fixed factors. (B) Bland–Altman comparison of MVC measurements obtained using the primary dynamometer (KinCom) and backup dynamometer (Biodex) in an independent cohort of volunteers (n=8). The mean bias was +10.9%, with the 95% limits of agreement shown in the figure.

Supplementary material 7: Fiber-type distribution and muscle fiber morphology

Immunofluorescence analyses of muscle fiber morphology and fiber-type distribution in muscle biopsies (n=54). (A) Type I muscle fiber cross-sectional area (fCSA). (B) Type I fiber shape factor index (SFI). (C) Fiber-type distribution expressed as the proportion of total fibers. (D) Baseline comparison of NCAM-positive fibers between younger and older participants. Data in panels A–C are shown as mean ± SD and were analyzed using mixed-effects models with Time, Age, and Group as fixed factors. Data in panel D are shown as median with individual values and were analyzed using a Mann–Whitney U test. Significance is indicated as ****p<0.0001.

Supplementary material 8: RNA sequencing analyses

(A) Hierarchically clustered sample-to-sample correlation heatmap showing separation primarily by Leg and secondarily by Age. (B) Principal component analysis of normalized gene expression using PC1 and PC2, with samples colored by Age, illustrating separation between younger and older participants primarily along PC2. (C) Gene Ontology cellular-component enrichment analysis of genes downregulated in the immobilized leg, highlighting mitochondrial gene sets. (D) Differential expression analysis of the effect of Leg restricted to the No-train group (FDR<0.05), showing an immobilization-induced transcriptional response comparable to that observed in the full cohort. (E) Differential expression analysis of the effect of Age restricted to control-leg biopsies (FDR<0.05), showing an ageing-associated transcriptional pattern comparable to that observed when both legs were included. (F) Correlations between WGCNA module eigengenes and Age, Leg, and Group.

**Supplementary Material 9: Training program used during prehabilitation and rehabilitation**

Number of sets, repetition ranges, and exercise intensity expressed as a percentage of one-repetition maximum.

**Supplementary Material 10: qPCR primers**

**Supplementary Material 11: Antibodies and reagents used for immunofluorescence analyses**

