## Supplemental material for "Prehabilitative Exercise Mitigates Immobilization-Induced Atrophy Through Muscle Reserve Rather Than Resistance to Disuse: The PRE-EX Randomized Controlled Trial"

### Supplemental material 1

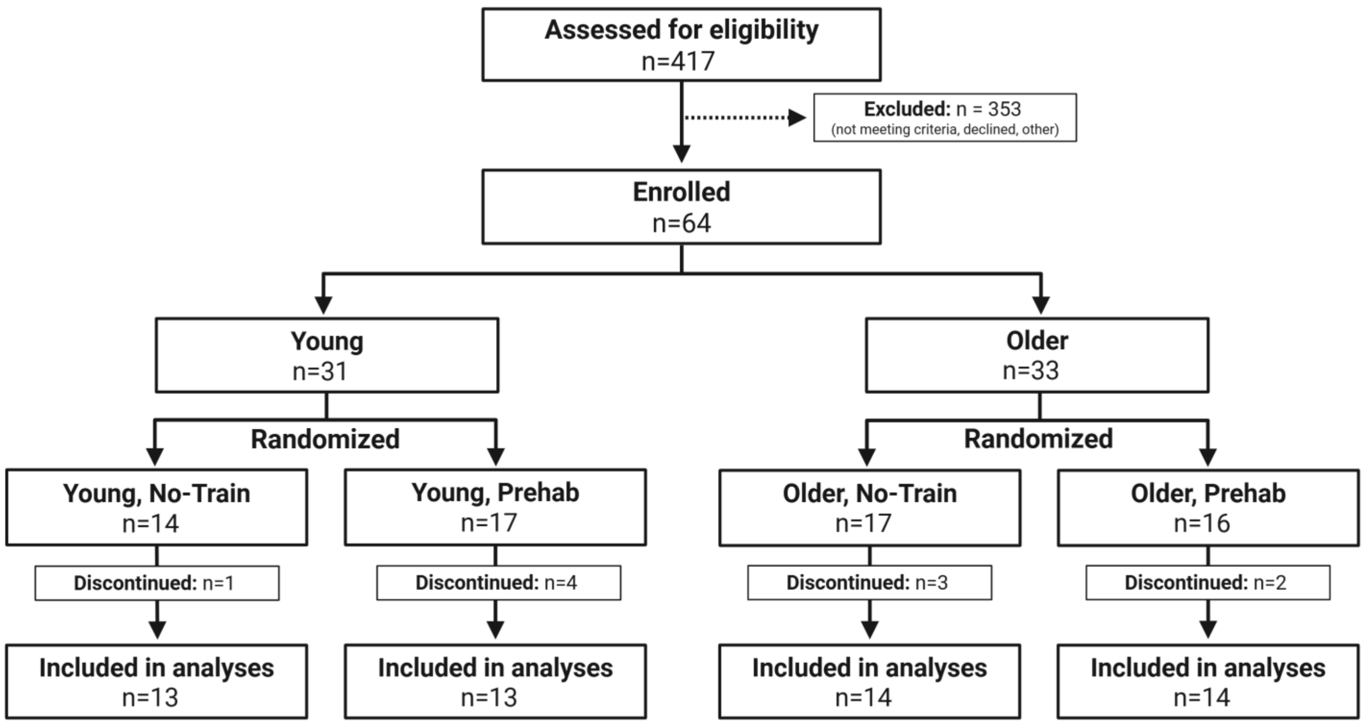

### Supplemental material 2

| | Young (n=26) | Older (n=28) | p value |
|---|---|---|---|
| Quadriceps CSA ($cm^2$) | 56±9 | 43±6 | **<0.0001** |
| MVC (Nm)[1] | 183±46 | 121±28 | **<0.0001** |
| Specific strength ($Nm/cm^2$)[1] | 3.36±0.60 | 2.87±0.57 | **<0.05** |
| Type I fCSA ($\mu m^2$) | 3634±599 | 3412±530 | 0.1545 |
| Type II fCSA ($\mu m^2$) | 3404±902 | 2187±524 | **<0.0001** |
| Type I SFI | 1.31±0.04 | 1.32±0.03 | 0.0631 |
| Type II SFI | 1.36±0.05 | 1.41±0.05 | **<0.001** |
| Type I fibers (%) | 61.1±13.9 | 54.7±14.4 | 0.1051 |

### Supplemental material 3

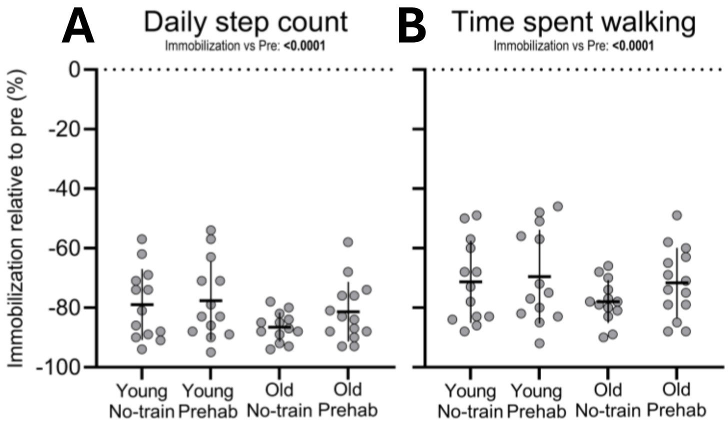

### Supplemental material 4

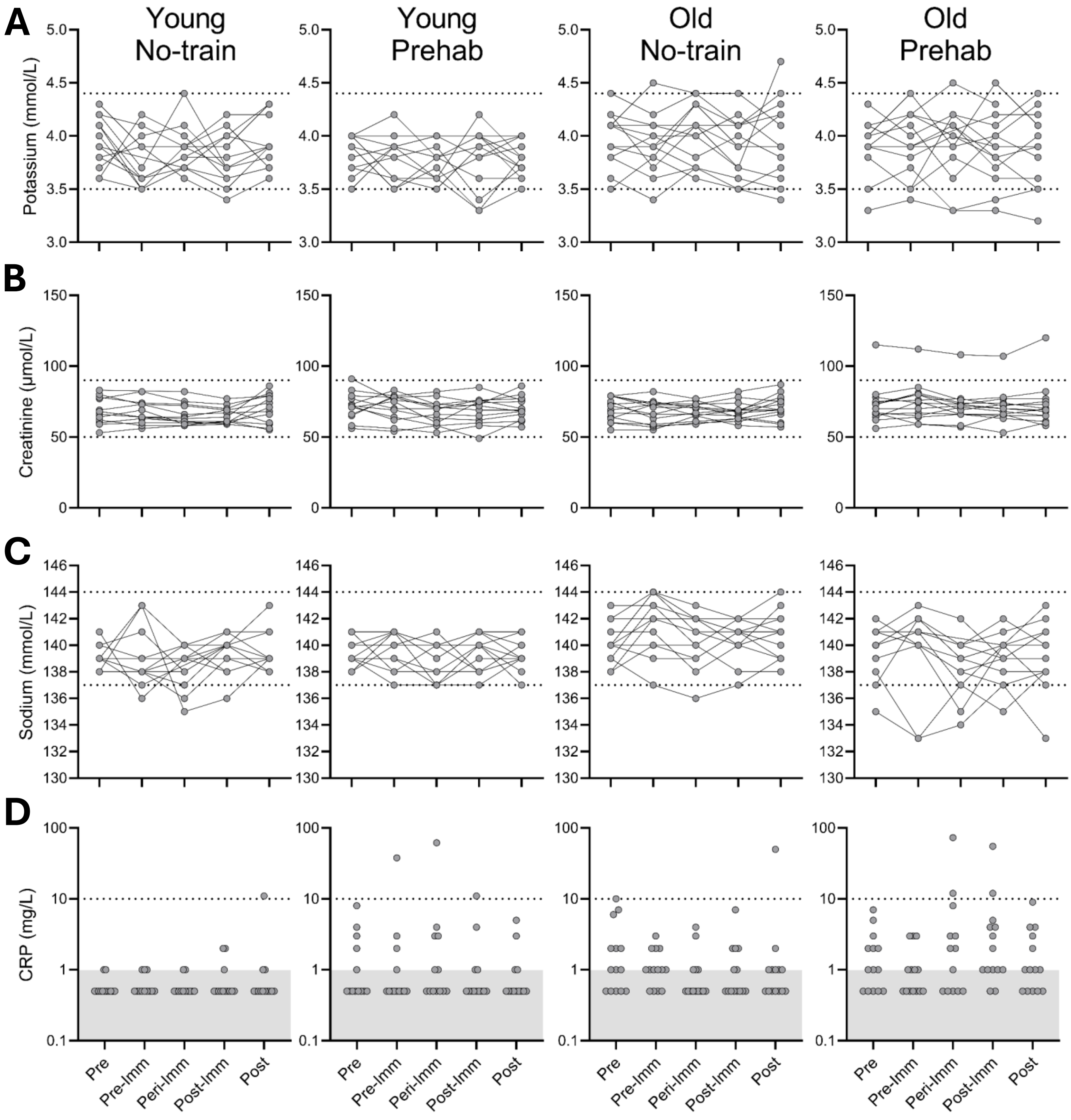

Supplemental material 5

Placeholder

### Supplemental material 6

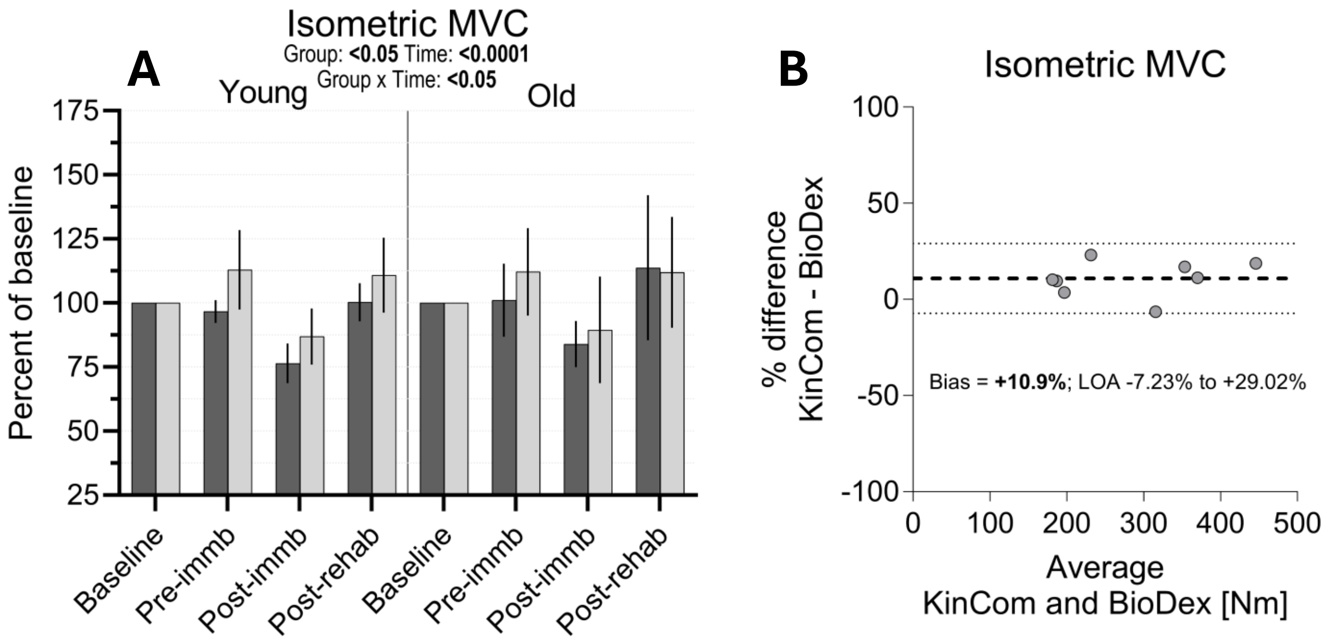

### Supplemental material 7

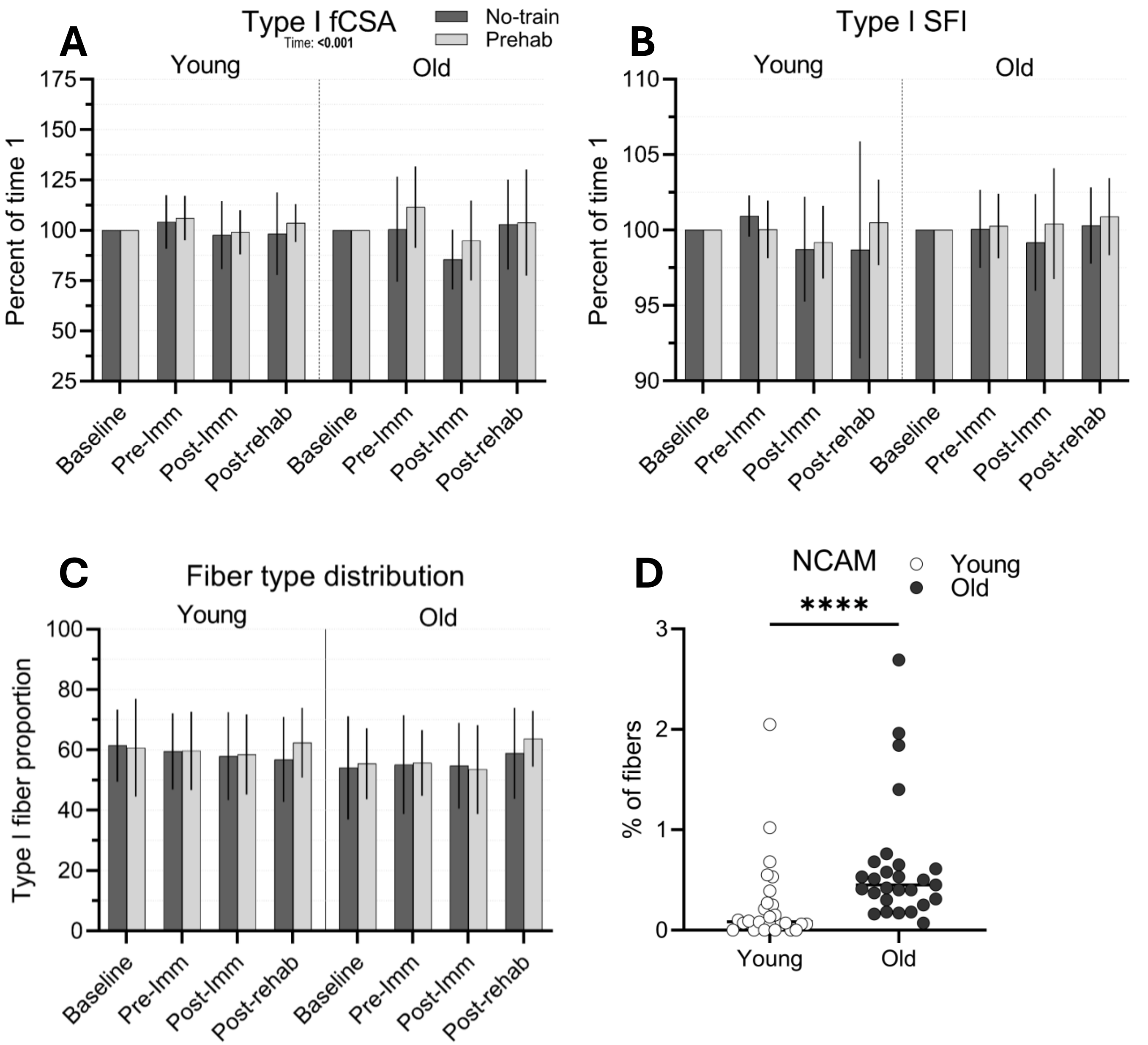

### Supplemental material 8

A

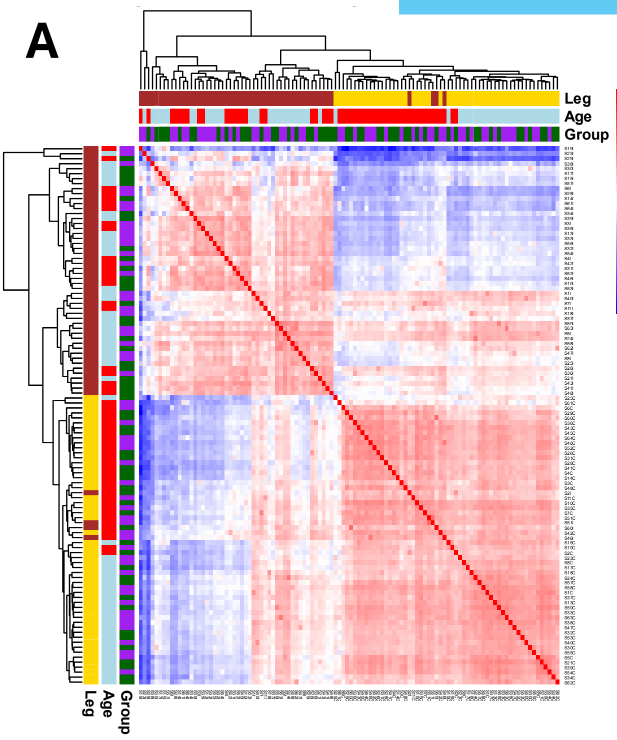


B

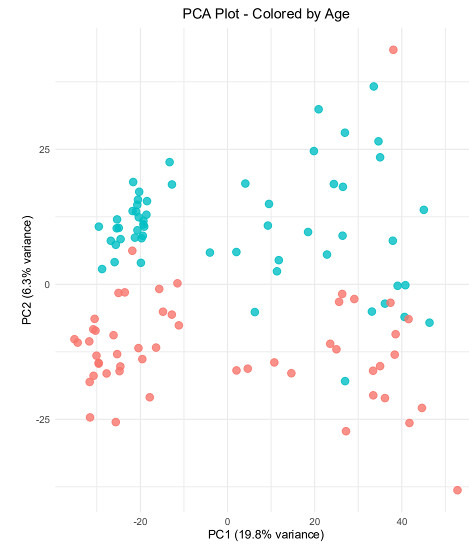


C

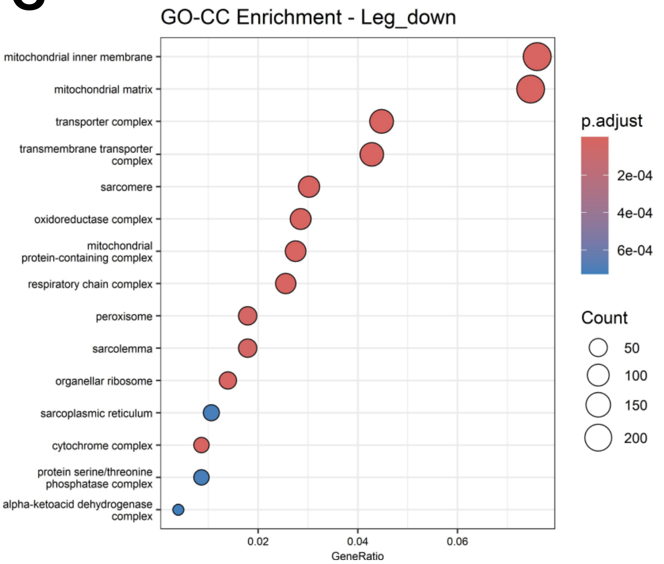


D

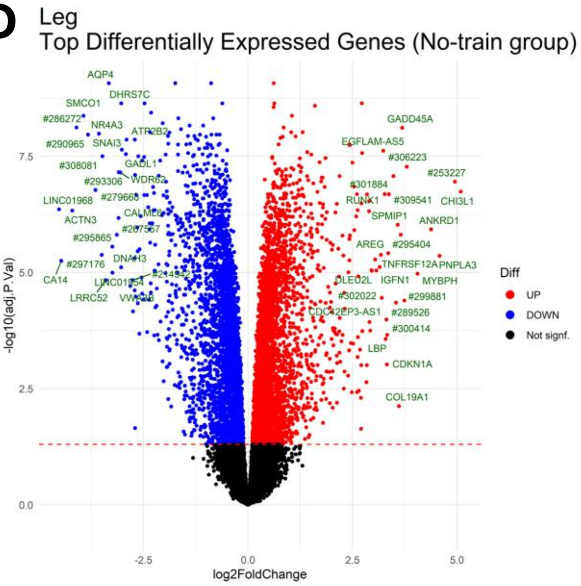


E

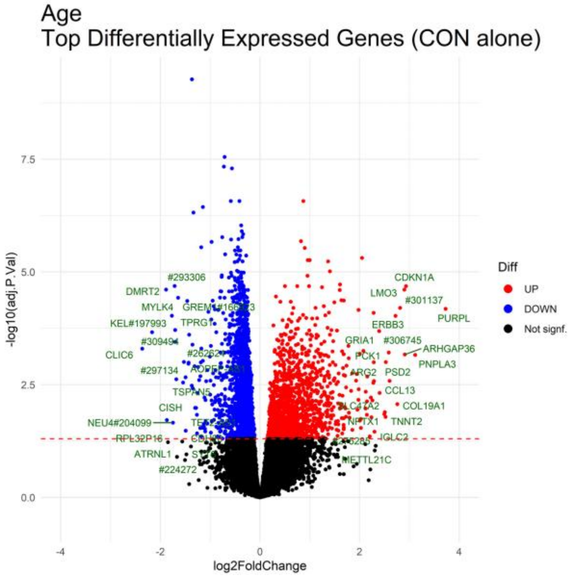


F

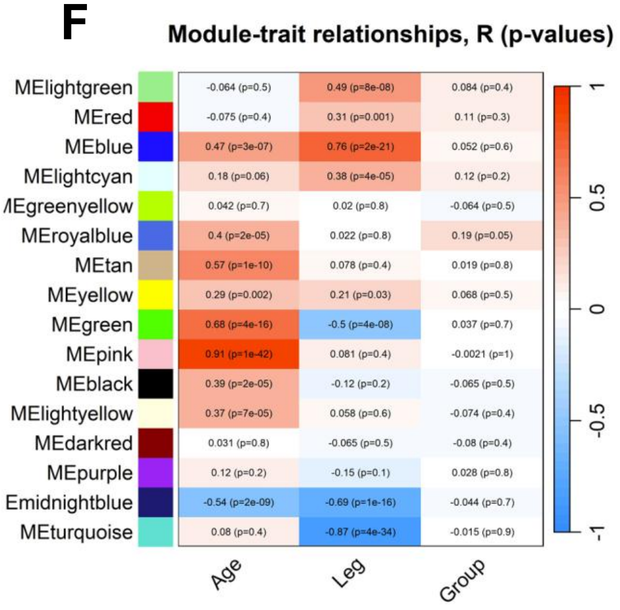

### Supplemental material 9

| Session | 1 | 2 | 3 | 4 | 5 | 6 | 7 | 8 | 9 | 10 |
|---|---|---|---|---|---|---|---|---|---|---|
| Leg press | | | | | | | | | | |
| Sets | 3 | 3 | 4 | 4 | 4 | 5 | 5 | 5 | 5 | 5 |
| Repetitions | 12-15 | 12-15 | 10-12 | 10-12 | 10-12 | 8-10 | 8-10 | 8-10 | 6-8 | 6-8 |
| Intensity (% of 1RM) | 60% | 60% | 69% | 69% | 69% | 74% | 74% | 74% | 79% | 79% |
| Knee extension | | | | | | | | | | |
| Sets | 3 | 3 | 4 | 4 | 4 | 5 | 5 | 5 | 5 | 5 |
| Repetitions | 12-15 | 12-15 | 10-12 | 10-12 | 10-12 | 8-10 | 8-10 | 8-10 | 6-8 | 6-8 |
| Intensity (% of 1RM) | 60% | 60% | 69% | 69% | 69% | 74% | 74% | 74% | 79% | 79% |
| Leg curl, pulldown, shoulder press | | | | | | | | | | |
| Sets | 3 | 3 | 3 | 3 | 3 | 4 | 4 | 4 | 4 | 4 |
| Repetitions | 12-15 | 12-15 | 12-15 | 12-15 | 12-15 | 10-12 | 10-12 | 10-12 | 10-12 | 10-12 |
| Intensity (% of 1RM) | 60% | 60% | 60% | 60% | 60% | 69% | 69% | 69% | 69% | 69% |

### Supplemental material 10

| mRNA | Gene name | Genbank | Sense | Antisense |
|---|---|---|---|---|
| RPLP0 | RPLP0 | NM_053275.3 | GGAAACTCTGCATTCTCGCTTCCT | CCAGGACTCGTTTGTACCCGTTG |
| GAPDH | GAPDH | NM_002046.4 | CCTCCTGCACCACCAACTGCTT | GAGGGGCCATCCACAGTCTTCT |
| AChR α1 | CHRNA1 | NM_000079.3 | GCAGAGACCATGAAGTCAGACCAGGAG | CCGATGATGCAAACAAGCATGAA |
| AChR δ | CHRND | NM_000751.2 | CAGCTGTGGATGGGGCAAAC | GCCACTCGGTTCCAGCTGTCTT |

### Supplemental material 11

#### PRIMARY ANTIBODIES

| Antibody | Host | Company | Cat. no. | Concentration | RRID | Assay |
|---|---|---|---|---|---|---|
| MyHC I, IgG2b | Mouse | DSHB | BA.D5 | 1:100 | AB_2235587 | Muscle fiber morphology |
| MyHC IIa, IgG1 | Mouse | DSHB | SC-71 | 1:100 | AB_2147165 | Muscle fiber morphology |
| Desmin | Rabbit | Abcam | AB32362 | 1:1000 | AB_731901 | Muscle cell culture |
| Myogenin, IgG1 | Mouse | DSHB | F5D | 1:50 | AB_2146602 | Muscle cell culture |
| Ki67, IgG2b | Mouse | Abcam | ab238020 | 1:1000 | AB_3076661 | Muscle cell culture |
| Tau1 | Rabbit | GeneTex | GTX130462 | 1:500 | AB_2886280 | Motor neuron cell culture |
| ChAT | Goat | Millipore | AB144p | 1:100 | AB_11212924 | Motor neuron cell culture |
| NCAM, IgG1 | Mouse | Becton Dickinson | 347740 | 1:50 | AB_400345 | Denervated fibers |
| Dystrophin, IgG2b | Mouse | Sigma-Aldrich | D8168 | 1:500 | AB_259245 | Denervated fibers |

#### SECONDARY ANTIBODIES

| Antibody | Host | Company | Cat. no. | Concentration | RRID | Assay |
|---|---|---|---|---|---|---|
| Anti-rabbit, IgG, 568 | Goat | Invitrogen | A-11036 | 1:200 | AB_10563566 | Muscle cell culture |
| Anti-mouse, IgG, 488 | Goat | Invitrogen | A-11029 | 1:200 | AB_2534088 | Muscle cell culture |
| Anti-goat, IgG, 568 | Donkey | Invitrogen | A-11057 | 1:200 | AB_2534104 | Motor neuron cell culture |
| Anti-rabbit, IgG, 488 | Donkey | Jackson ImmunoResearch Labs | 711-545-152 | 1:100 | AB_2313584 | Motor neuron cell culture |
| Anti-mouse, IgG2b, 488 | Goat | Invitrogen | A-21141 | 1:250 | AB_141626 | Denervated fibers |
| Anti-mouse, IgG2b, 488 | Goat | Invitrogen | A-21141 | 1:500 | AB_141626 | Muscle fiber morphology |
| Anti-mouse, IgG1, 568 | Goat | Invitrogen | A-21124 | 1:250 | AB_2535766 | Denervated fibers |
| Anti-mouse, IgG1, 568 | Goat | Invitrogen | A-21124 | 1:500 | AB_2535766 | Muscle fiber morphology |

#### DYES

| Antibody | Host | Company | Cat. no. | Concentration | RRID | Assay |
|---|---|---|---|---|---|---|
| Wheat Germ Agglutinin | N/A | Invitrogen | W32465 | 1:500 | N/A | Muscle fiber morphology |
